# Diagnosis provision by young people’s mental health services: a comparison with epidemiological data

**DOI:** 10.64898/2026.05.28.26354156

**Authors:** Stephanie J Lewis, Alan J Meehan, Mia Akiba, Louise Arseneault, Sarah Byford, Avshalom Caspi, Bruce R Clark, Johnny Downs, Tamsin J Ford, Helen L Fisher, Karestan C Koenen, Terrie E Moffitt, Joanne B Newbury, Candice L Odgers, Megan Pritchard, Emily Simonoff, Andrea Danese

## Abstract

**Background:** Little is known about the provision of diagnoses to young people with mental health disorders. We investigated diagnosis provision by NHS mental health services, focusing on 17-year-olds in South London between 2009-2024, and compared with estimated disorder prevalence.

**Methods:** To examine diagnosis provision in the population, we extracted diagnosis data from records of the NHS mental healthcare provider serving South London, using the Maudsley Biomedical Research Centre Clinical Record Interactive Search application; we then compared these data with the corresponding population size, obtained from the Office for National Statistics. To assess diagnosis provision in those with mental health disorders, we compared diagnosis data with the number of young people estimated to have met criteria for a disorder, derived from epidemiological interview data collected in the Environmental Risk (E-Risk) Longitudinal Twin Study and weighted according to characteristics of 17-year-old South Londoners. To assess diagnosis provision in those with mental health disorders within health services, we compared diagnosis data with the number estimated to have met criteria for a disorder and used any health service for their mental health, again derived from weighted E-Risk Study data.

**Findings:** Of 17-year-olds from South London in 2009-2024, 4.0% (n=8,958/223,404) had a diagnosis in mental health records during the previous year. This diagnosis provision covered <1 in 16 of those estimated to have had a mental health disorder, and <1 in 4 of those estimated to have also used health services. Diagnosis provision was lower in girls than boys and in young people with Black/Asian/Mixed/Other ethnicity than those with White ethnicity, in those estimated to have had a mental health disorder and used health services.

**Interpretation:** These findings demonstrate gaps and biases in mental health diagnosis provision for young people, including within health services, and reveal the imperative need to strengthen young people’s mental healthcare.

**Funding:** National Institute for Health Research, Medical Research Council, National Institute of Child Health and Development, Jacobs Foundation, National Society for Prevention of Cruelty to Children, Economic and Social Research Council, Prudence Trust, Wellcome Trust

**RESEARCH IN CONTEXT:** *Evidence before this study:* Epidemiological research has found that less than half of young people with a mental health disorder report seeing a health professional for their symptoms and only a quarter to a third have seen a mental health specialist, and these findings are fairly consistent across high-income countries where this research has been conducted. For young people who see health professionals, there are likely to be barriers to the recognition and treatment of mental health disorders. We focused on the recognition of mental health disorders in young people, which is operationalised in clinical practice as provision of diagnoses. To identify studies examining mental health diagnosis provision in young people accessing health services, we searched MEDLINE with the terms “diagnos*” AND (“mental” OR “psychiatr*” OR [various terms for individual disorders]) AND (“young” OR “youth*” OR “child*” OR “adolescen*” OR “paedatric*” OR “pedatric*” OR “juvenile”) AND (“clinical” OR “healthcare” OR “health care” OR “service*”). This search was supplemented by reviewing reference lists and forward citations of relevant articles. We identified several studies that found diagnosis provision varied by sociodemographic characteristics and has increased over the past two decades in young people across multiple countries for several disorders, including depression, anxiety disorders, eating disorders, autism, and attention-deficit/hyperactivity disorder (ADHD). Only a small number of studies investigated diagnosis provision within young people who met criteria for a disorder. In the Avon Longitudinal Study of Parents and Children in the UK, of 18-year-olds who met criteria for depression, only 7.0% had a diagnosis of depression documented in their primary care records. In the Child and Adolescent Twin Study in Sweden, of 9-year-olds who met criteria for ADHD, only 18.5% of boys and 12.1% of girls had a diagnosis of ADHD noted in their health records. Within an Irish child and adolescent mental health service, of 12-15-year-olds who met criteria for depressive disorder, only 28.4% received a depressive disorder diagnosis after their usual clinical assessment. These studies suggest large gaps in diagnosis provision, including within health services, and highlight possible bias by sociodemographic characteristics. A better understanding of this topic is needed to enable more effective service planning, commissioning, and policymaking.

*Added value of this study:* This study investigated mental health diagnosis provision in 17-year-olds from South London. We examined diagnosis provision for several mental health disorders in NHS mental healthcare records. We compared these data with population and epidemiological data to calculate diagnosis provision rates in the general population, in those estimated to have met criteria for a disorder, and in those estimated to have also seen a health professional. We found that diagnosis provision substantially increased during our study period of 2009-2024, demonstrating an increase in the number of young people whose mental health needs were recognised in specialist services. However, we estimated that diagnoses were only provided to a small proportion of young people with a mental health disorder, including those within health services. Estimated diagnosis gaps were largest for those with generalised anxiety disorder and multiple disorders. We also found evidence of biases in diagnosis provision, based on gender, neighbourhood deprivation, and ethnicity.

*Implications of all the available evidence:* Barriers to mental healthcare access for young people should be reduced by policymakers and commissioners, including through investment in adequately staffed services with skilled clinicians, enabling more young people with mental health disorders to receive cost-effective evidence-based healthcare that has long-term benefits. Greater awareness among clinicians of the under-diagnosis and bias in diagnosis of young people’s mental health disorders, alongside strategies to address these problems, could improve young people’s mental healthcare. Innovative scalable interventions that can reach many more young people need to be developed, evaluated, and implemented by researchers. Prevention strategies are also required, including addressing risk factors for young people’s mental health disorders, and intervening early for those with symptoms before disorders develop, to reduce the future burden of mental health disorders in young people.

## INTRODUCTION

Mental health disorders cause substantial distress for affected young people and, if left untreated, can have long-term negative effects on their health, education, and employment.^1,2^ Importantly, several specialist interventions have been found to be clinically- and cost-effective, with lasting benefits for individuals and society.^3^ There is, therefore, a strong moral and economic case to ensure that such evidence-based treatments can be obtained by young people with mental health disorders. However, accessing treatments can be challenging and requires several steps (Figure 1). Research and policy on healthcare provision have largely focused on early steps, such as help-seeking and service use.^4–8^ Understanding gaps in later steps, such as diagnosis provision and treatment access, is also crucial to improving healthcare for young people by highlighting targets for service-level interventions. Conversely, identifying over-diagnosis or over-treatment can inform strategies for more efficient use of available resources.

**Figure 1.**
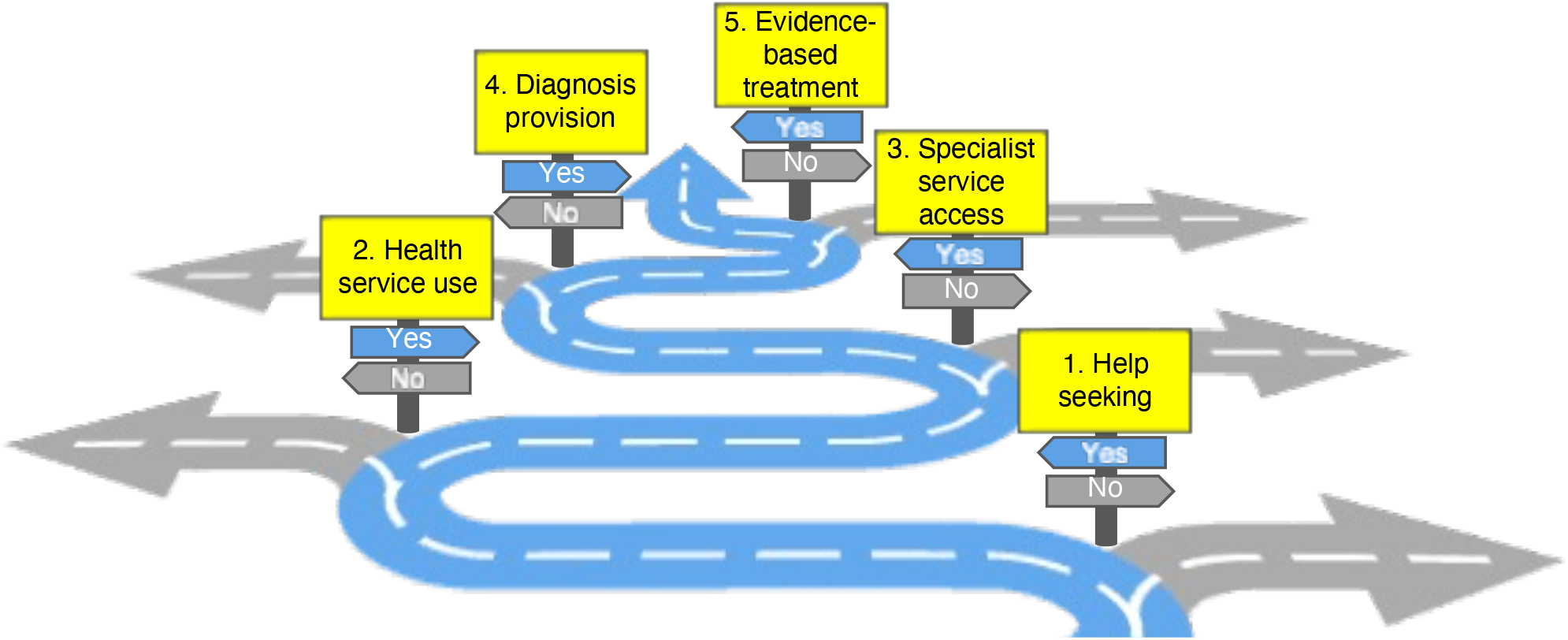
Route to evidence-based treatment. This figure illustrates the route to evidence-based treatment for most young people with mental health disorders, shown in blue, on which there are several junctions to be navigated, including the five summarised here: 1. First, the young person or their caregivers need to seek help from health professionals, sometimes with the support of others (e.g. teachers). 2. Then, they need to use those health services. The health professional needs to recognise that the young person has a likely mental health disorder and refer to services that can provide further care. The skills and ongoing supervision needed for recommended management of mental health disorders in young people are largely only available in specialist mental health services in the UK, which includes a range of teams from those offering low intensity interventions to those providing the most complex care (appendix p2-3). 3. Next, the young person needs to access these specialist services, which requires that the referral is accepted and that the young person attends. 4. Then, the mental health specialist must recognise and provide an accurate diagnosis of the disorder. This step is important because current research evidence and guidelines on treatment are disorder-specific. That is, treatments that have been found to be effective and are recommended for a particular disorder (e.g. attention-deficit/hyperactivity disorder) are not known to be effective or recommended for other disorders (e.g. depressive disorder). So, if a young person is misdiagnosed with another or no disorder, they will not receive treatments that have been found to be effective for their condition. 5. After diagnosis provision, evidence-based treatment can be offered if available, and delivered if the young person engages. Although this route may not be helpful for all young people with mental health disorders, evidence suggests that it is likely to be beneficial for most. Therefore, this management is recommended in healthcare guidelines, such as those produced by the UK National Institute for Health and Care Excellence (appendix p2-3). According to the NHS Constitution for England, such healthcare should be available to all who need it.

In this study, we investigated diagnosis provision, which indicates the recognition of a disorder by a clinician (Figure 1, Step 4). Diagnosis provision signifies the threshold for treatment in the UK, and guides the delivery of disorder-specific treatments. Studying three nested samples of 17-year-olds from South London (Figure 2), we examined diagnoses documented in NHS mental health records, compared with population and epidemiological data to calculate diagnosis provision rates, and tested for sociodemographic differences. We hypothesised that diagnosis provision does not meet need even within health services, and provision might be lower in some sociodemographic groups.

**Figure 2.**
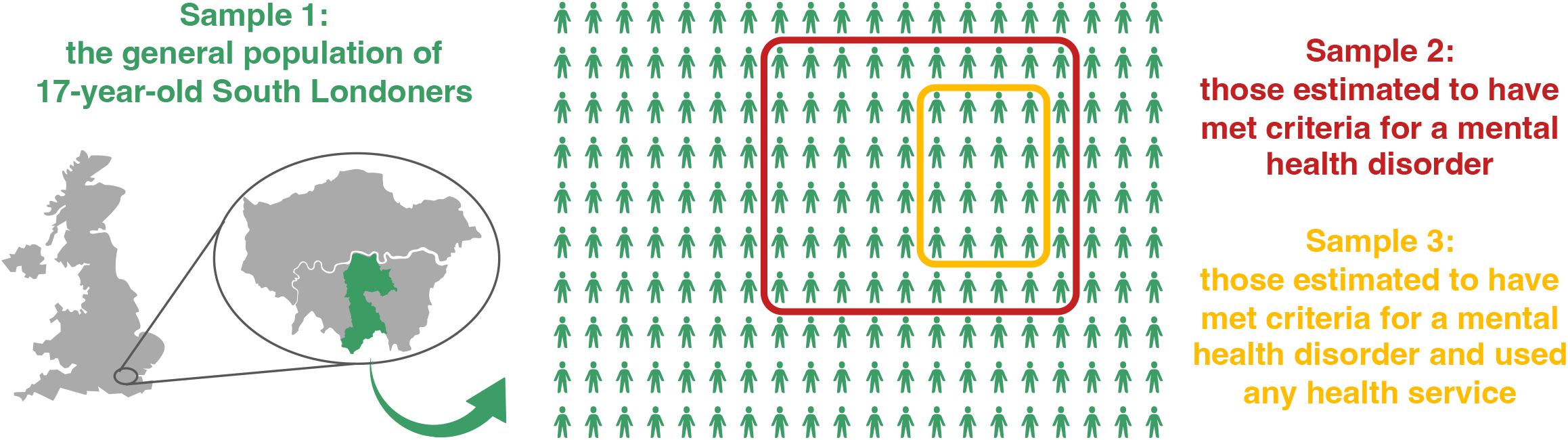
Nested samples studied. The figure illustrates the three nested samples studied: Sample 1 (green), the general population of all 17-year-olds living in South London (boroughs of Croydon, Lambeth, Lewisham, and Southwark in the UK) in 2009-2024; Sample 2 (red, within Sample 1), those estimated to have met criteria for a mental health disorder during the previous year; and Sample 3 (yellow, within Samples 1 and 2), those estimated to have also used any health service for help with mental health problems during that year. Sample 1 was identified from population census and administrative data, and Samples 2 and 3 were derived from epidemiological interview data weighted and applied to population data.

## METHODS

### Data source for mental health diagnosis provision

We obtained data about mental health diagnosis provision from South London and Maudsley NHS Foundation Trust, which is one of Europe’s largest mental healthcare providers, and has a robust infrastructure to process electronic health records for research, offering a unique data resource.^9^ This organisation is the sole provider of state-funded universal specialist mental healthcare for the population of four South London boroughs. This provider is responsible for the delivery of state-funded evidence-based treatments for children and adolescents with mental health disorders in this area. Other interventions, such as supportive counselling, are available from other services and may be appropriate for subthreshold symptoms, but are not recommended for the treatment of disorders (appendix p2-3).

We extracted data from these records via the Maudsley Biomedical Research Centre Case Register using the Clinical Record Interactive Search application.^9^ We selected data on young people from South London who were aged 17 (i.e., 17 ≤age<18) years on 30th June each year in 2009-2024 and had an active mental health diagnosis during the previous year. We chose age 17 years to capture those who received care from child and adolescent mental health services and to best fit with data for estimated prevalences of disorders and health service use (described below). We chose 30th June to align with data for population characteristics (described below). We extracted patients’ diagnosis data according to ICD-10 (listed in Figure 3, detailed in appendix p4), from structured fields which clinicians must complete before patients can be discharged. We also extracted patients’ sociodemographic data, including gender, ethnicity, and neighbourhood deprivation (Index of Multiple Deprivation national quintiles, based on their home address’ lower-layer super output area that comprises approximately 1,500 residents) at diagnosis.

**Figure 3.**
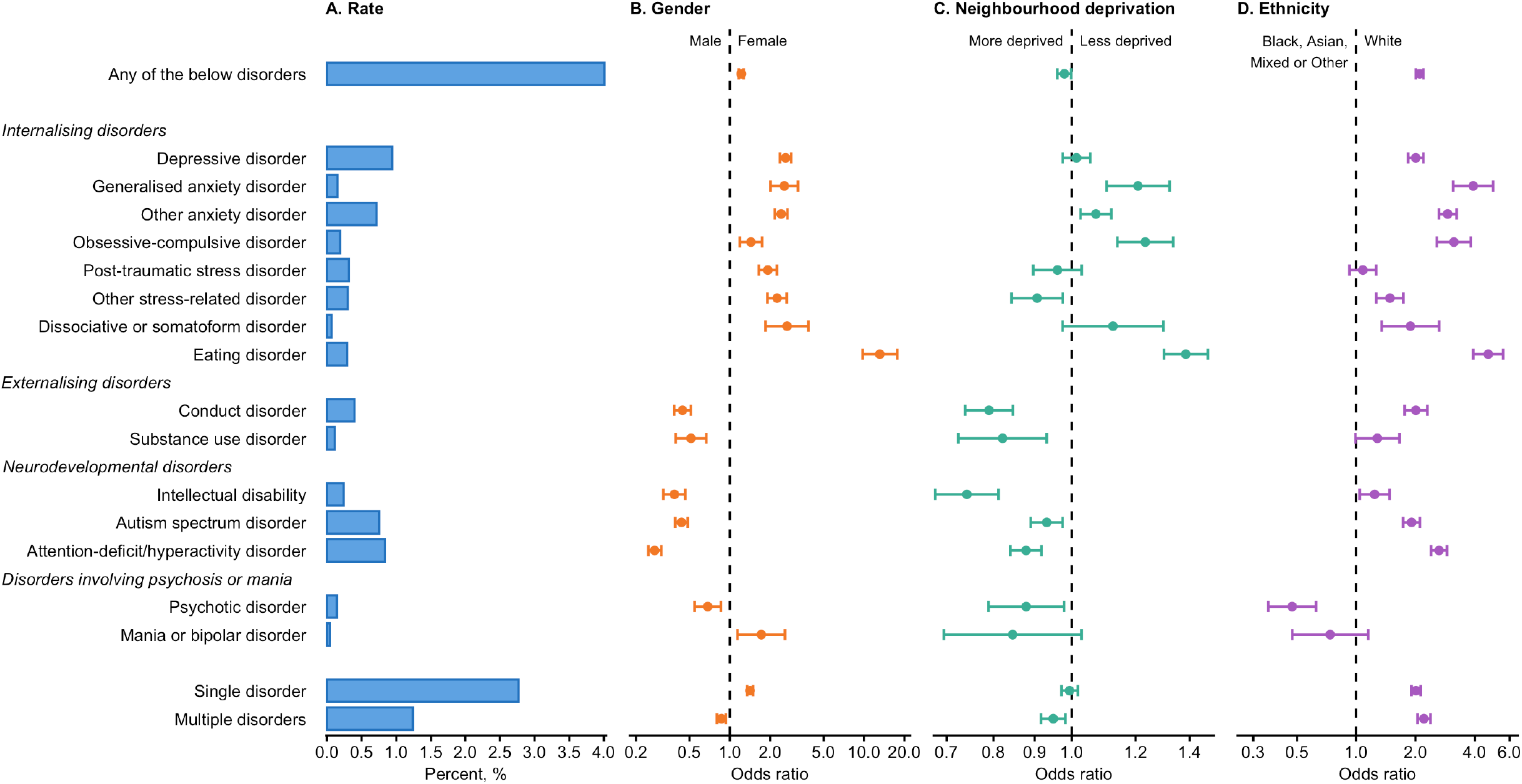
Mental health diagnosis provision in the general population of 17-year-olds from South London, Sample 1. The plot presents the rates of mental health diagnosis provision during the past year in 17-year-olds from South London in 2009-2024, based on diagnoses documented in NHS mental health records compared with population census and administrative data (Panel A). The plot also presents odds ratios and their 95% confidence intervals for associations of gender (female vs male), neighbourhood deprivation (Index of Multiple Deprivation national quintile; 1=most deprived, 5=least deprived), and ethnicity (White vs Black, Asian, Mixed, or Other ethnicity) with diagnosis provision in these young people (Panels B, C, and D respectively). For more details, see appendix p19-24.

### Data source for population characteristics

We next obtained the size of the general population to which these young people with diagnoses belonged. The UK Office for National Statistics determines population size as of 30th June annually, based on the previous census and subsequent administrative data. From this resource, we sought the number of 17-year-olds who lived in South London in 2009-2024 (Figure 2, Sample 1), stratified by sex (as a proxy for gender) and by neighbourhood deprivation (Index of Multiple Deprivation based on lower-layer super output areas). Additionally, we ascertained the ethnicity of the population by applying ethnicity ratios found in censuses.

### Data source for mental health disorders and health service use

We then obtained prevalence rates of mental health disorders and of health service use in this population. Because interviewing the entire population to obtain exact prevalence figures is not feasible, we derived estimates from epidemiological research. To identify a comparable epidemiological study, we systematically reviewed existing cohorts and identified samples representative of British young people, where data could be weighted to the sociodemographic characteristics of South London, and where measures involved interviews to assess past-year mental health disorders and health service use. We determined that the study that best fit our prespecified requirements was the Environmental Risk (E-Risk) Longitudinal Twin Study (appendix p7-13).

The E-Risk Study has tracked the development of a birth cohort of 2,232 children born in England and Wales in 1994-95, which is representative of the UK population.^10^ At 18 years old in 2012-2013, 2,066 participants (93% of the original sample) were interviewed during home visits. Assessment of mental health disorders (depressive disorder, generalised anxiety disorder, post-traumatic stress disorder [PTSD], conduct disorder, and attention-deficit/hyperactivity disorder [ADHD]) was undertaken using the Diagnostic Interview Schedule for DSM-IV or adapted structured interviews to ascertain DSM-5 diagnostic criteria, including impairment, in the past year (thus for the period when the participants were aged 17 years). Such research interviews provide prevalence estimates that are similar to estimates based on clinical interviews,^11,12^ but are more feasible in large-scale epidemiological studies. Health service use for mental health problems in the past year, including primary care and specialist services, was also assessed by interview.

We used this E-Risk Study data to estimate prevalence rates of mental health disorders and health service use weighted according to sex and neighbourhood deprivation distributions in 17-year-old South Londoners. We additionally undertook sensitivity analyses that also weighted according to ethnicity distributions. We then applied these rates to population data to calculate the estimated number of 17-year-olds from South London who would have met criteria for a disorder during the past year in 2009-2024 (Figure 2, Sample 2), and the estimated number who would have also used health services (Figure 2, Sample 3), by sex (as a proxy for gender), neighbourhood deprivation, and ethnicity.

### Statistical analysis

We investigated the provision of mental health diagnoses in our three nested samples (described above and in Figure 2). To calculate the rate of mental health diagnosis provision in each sample, we determined the number of 17-year-old South Londoners who had a diagnosis in mental health records during the previous year, and divided it by the number of young people in each sample. To identify differences by sociodemographic characteristics, we tested for associations of gender, neighbourhood deprivation, and ethnicity with diagnosis provision using logistic regression.

Further details on our methods are provided in appendix p4-18.

## RESULTS

### 1. Mental health diagnosis provision in the general population of 17-year-olds from South London, Sample 1

On analysing NHS mental health records of 17-year-olds from South London in 2009-2024, we found that 8,958 had a diagnosis of the mental health disorders listed in Figure 3 during the previous year. According to the Office for National Statistics, there were 223,404 17-year-olds living in South London in 2009-2024 (Sample 1). By dividing these numbers, we calculated that the rate of diagnosis provision in the previous year was 4.0% (n=8,958/223,404) in Sample 1. The most common diagnoses were depressive disorder (0.9%, n=2,105/223,404), ADHD (0.8%, n=1,875/223,404), and autism spectrum disorder (0.8%, n=1,686/223,404). Of those with a diagnosis, 30.9% (n=2,771/8,958) were diagnosed with multiple disorders (Figure 3, Panel A). When considered by year, diagnosis provision rate increased 1.7-fold over the 16-year period, from 2.6% (n=355/13,777) in 2009 to 4.4% (n=674/15,191) in 2024 (appendix p20-21).

Sociodemographic characteristics were associated with diagnosis provision in Sample 1. Girls were more likely to be diagnosed with any disorder, internalising disorders, and mania or bipolar disorder than boys; whereas boys were more likely to be diagnosed with externalising, neurodevelopmental, and psychotic disorders than girls (Figure 3, Panel B). Young people living in less deprived neighbourhoods were more likely to be diagnosed with generalised anxiety, other anxiety, obsessive-compulsive, and eating disorders than those from more deprived neighbourhoods; whereas those living in more deprived neighbourhoods were more likely to be diagnosed with any disorder, other stress-related disorder, externalising disorders, neurodevelopmental disorders, and psychotic disorder than those from less deprived neighbourhoods (Figure 3, Panel C). Those with White ethnicity were more likely to be diagnosed with most disorders than those with Black, Asian, Mixed, or Other ethnicities; whereas those with these ethnicities were more likely to be diagnosed with a psychotic disorder than their White peers (Figure 3, Panel D). These results reflect sociodemographic differences in population prevalence of disorders, health service use, and/or diagnosis provision within services.

### 2. Mental health diagnosis provision in those estimated to have met criteria for a disorder, Sample 2

In subsequent analyses, we focused on five disorders assessed in the E-Risk Study: depressive disorder, generalised anxiety disorder, PTSD, conduct disorder, and ADHD. On analysing NHS mental health records of 17-year-olds from South London in 2009-2024, we found that 5,132 had a diagnosis of any of these disorders during the previous year. Based on E-Risk Study interview data weighted according to sex and neighbourhood deprivation distributions of 17-year-old South Londoners in 2009-2024, we estimated that 36.3% of our population would have met criteria for any of these disorders during the previous year. By applying this disorder prevalence to population data (n=223,404), we estimated that 81,050 17-year-olds from South London in 2009-2024 would have met criteria for any of these disorders during the previous year (Sample 2). By dividing these numbers, we calculated that the rate of diagnosis provision in the past year was 6.3% (n=5,132/81,050) in Sample 2. In other words, 6.3% of those estimated to have met criteria for any of these mental health disorders received a diagnosis from NHS mental health services. Young people estimated to have met criteria for ADHD were most likely to receive a diagnosis (9.8%, n=1,875/19,102), whereas those estimated to have had generalised anxiety disorder were least likely to receive a diagnosis (2.0%, n=340/17,131) (Figure 4, Panel A). Depressive disorder accounted for the highest number of estimated undiagnosed cases because of its high population prevalence (diagnosed n=2,105/45,755) (Figure 6). Regarding the number of disorders diagnosed, substantially fewer young people estimated to have met criteria for multiple disorders were diagnosed with more than one disorder (2.5%, n=748/29,617), compared with those estimated to have a single disorder who were diagnosed with one disorder (8.5%, n=4,384/51,433). Sensitivity analyses weighting according to ethnicity, as well as sex and neighbourhood deprivation, found similar results (appendix p26).

**Figure 4.**
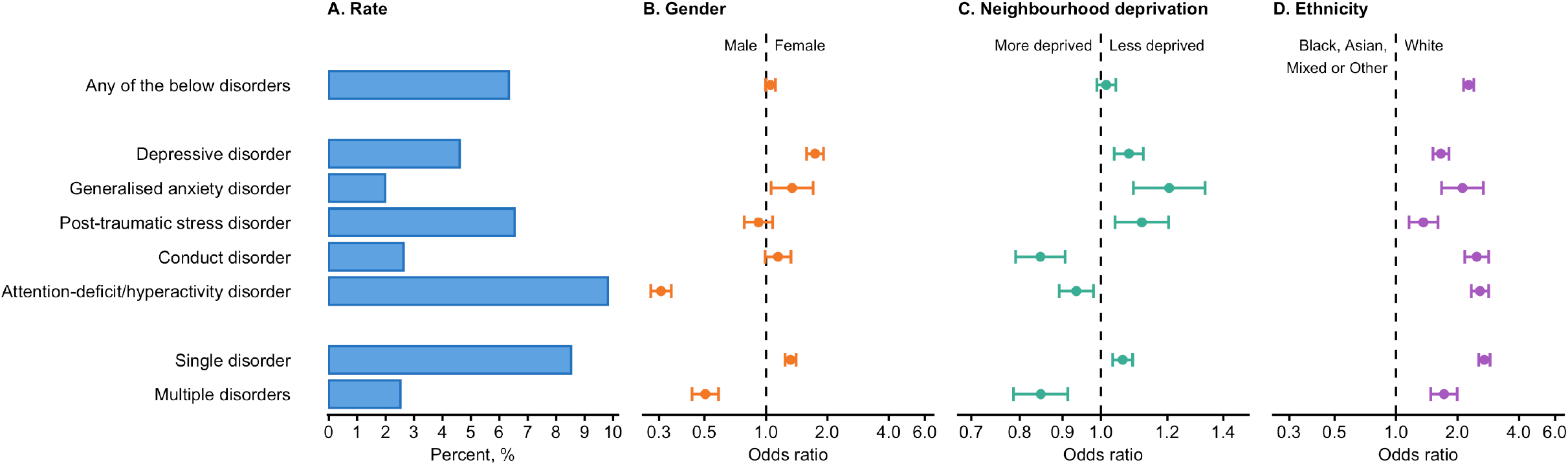
Mental health diagnosis provision in those estimated to have met criteria for a disorder, Sample 2. The plot presents the rates of mental health diagnosis provision in 17-year-olds from South London in 2009-2020 who were estimated to have met criteria for the disorder during the past year, based on diagnoses documented in NHS mental health records compared with epidemiological interview data weighted and applied to population data (Panel A). The plot also presents odds ratios and their 95% confidence intervals for associations of gender (female vs male), neighbourhood deprivation (Index of Multiple Deprivation national quintile; 1=most deprived, 5=least deprived), and ethnicity (White vs Black, Asian, Mixed, or Other ethnicity) with diagnosis provision in these young people (Panels B, C, and D respectively). For more details, see appendix p25-29.

Sociodemographic characteristics were associated with diagnosis provision in Sample 2. Boys estimated to have had depressive or generalised anxiety disorders were less likely to be diagnosed than girls with these disorders; whereas girls estimated to have had ADHD were less likely to be diagnosed than boys with this disorder (Figure 4, Panel B). Young people living in more deprived neighbourhoods who were estimated to have had depressive disorder, generalised anxiety disorder, and PTSD were less likely to be diagnosed than those from less deprived neighbourhoods with these disorders; whereas those living in less deprived neighbourhoods who were estimated to have had conduct disorder and ADHD were less likely to be diagnosed than those from more deprived neighbourhoods with these disorders (Figure 4, Panel C). Those with Black, Asian, Mixed, or Other ethnicity who were estimated to have had any of the five disorders were less likely to be diagnosed than their White peers with these disorders (Figure 4, Panel D). These results reflect sociodemographic differences in health service use and/or diagnosis provision within services (after accounting for disorder prevalence).

### 3. Mental health diagnosis provision in those estimated to have met criteria for a disorder and used health services, Sample 3

Based on E-Risk Study interview data weighted according to sex and neighbourhood deprivation distributions of 17-year-old South Londoners in 2009-2024, we estimated that 9.7% of our population would have met criteria for any of the five disorders and used health services for help with mental health problems during the previous year. By applying this disorder and health service use prevalence to population data, we estimated that 21,598 17-year-old South Londoners in 2009-2024 would have met criteria for these disorders and used health services during the previous year (Sample 3). We compared this number to the 5,132 17-year-old South Londoners in 2009-2024 who had a diagnosis of these disorders in NHS mental health records during the previous year, and calculated that the rate of diagnosis provision in the past year was 23.8% (n=5,132/21,598) in Sample 3. In other words, 23.8% of those estimated to have met criteria for any of these disorders and to have used health services received a diagnosis from NHS mental health services. Young people estimated to have met criteria for ADHD and to have used health services were most likely to receive a diagnosis (34.4%, n=1,875/5,454), whereas those estimated to have had generalised anxiety disorder and used health services were least likely to receive a diagnosis (3.9%, n=340/8,778) (Figure 5, Panel A). But again, depressive disorder accounted for the highest number of estimated undiagnosed cases in service users because of its high prevalence (diagnosed n=2,105/17,243) (Figure 6). Regarding the number of disorders diagnosed, far fewer young people estimated to have met criteria for multiple disorders and to have used health services were diagnosed with more than one disorder (6.0%, n=748/12,379), compared with those estimated to have had a single disorder and to have used services who were diagnosed with one disorder (47.6%, n=4,384/9,219). Sensitivity analyses weighting according to ethnicity, as well as sex and neighbourhood deprivation, found similar results (appendix p31).

**Figure 5.**
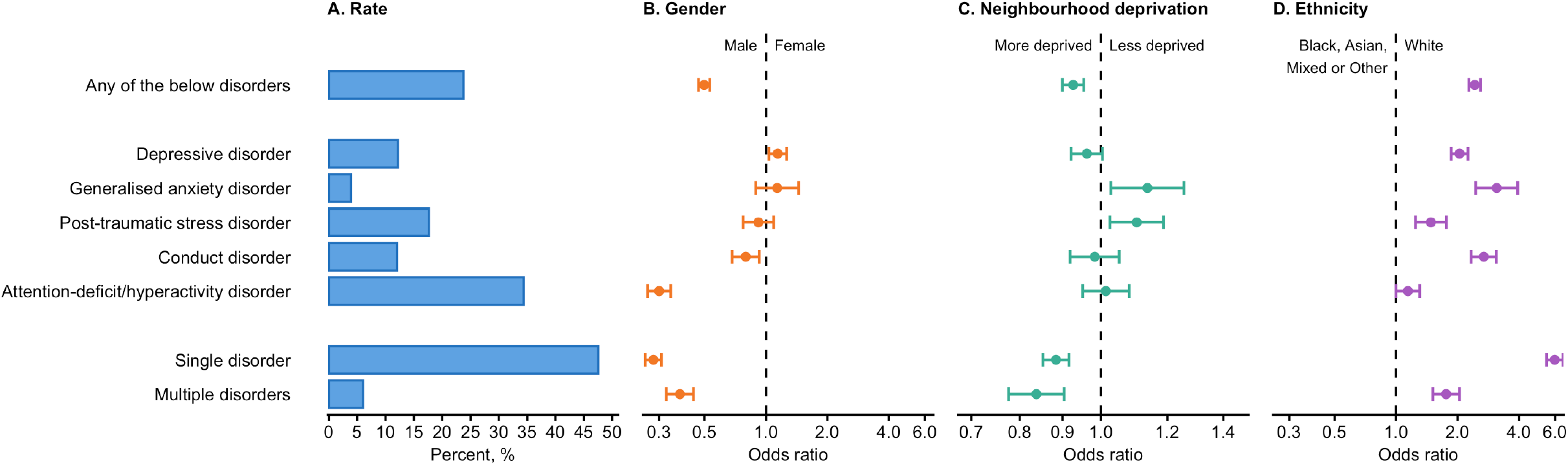
Mental health diagnosis provision in those estimated to have met criteria for a disorder and used health services, Sample 3. The plot presents the rates of mental health diagnosis provision in 17-year-olds from South London in 2009-2024 who were estimated to have met criteria for the disorder and to have used health services during the past year, based on diagnoses documented in NHS mental health records compared with epidemiological interview data weighted and applied to population data (Panel A). The plot also presents odds ratios and their 95% confidence intervals for associations of gender (female vs male), neighbourhood deprivation (Index of Multiple Deprivation national quintile; 1=most deprived, 5=least deprived), and ethnicity (White vs Black, Asian, Mixed, or Other ethnicity) with diagnosis provision in these young people (Panels B, C, and D respectively). For more details, see appendix p30-34.

**Figure 6.**
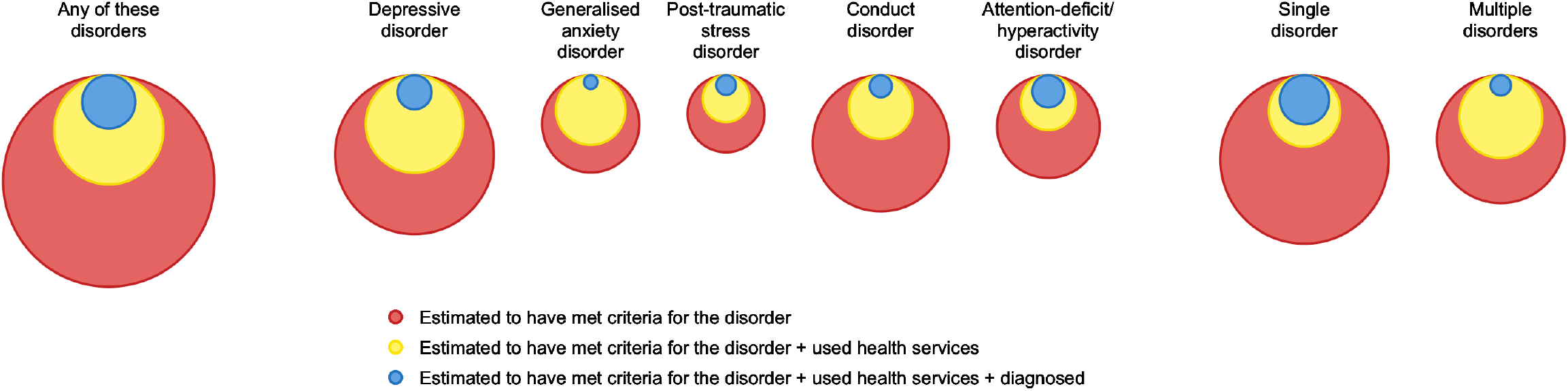
Mental health disorder, health service use, and diagnosis provision in the general population. The plot reflects the estimated prevalence of disorder (red circle area), disorder and health service use (yellow circle area), and rate of NHS mental health service diagnosis provision (blue circle area) during the past year in 17-year-olds from South London in 2009-2024.

Sociodemographic characteristics were associated with diagnosis provision in Sample 3. Boys estimated to have had depression and to have used health services were less likely to be diagnosed than girls with this disorder who used services; whereas girls estimated to have had any disorder, conduct disorder, or ADHD and to have used health services were less likely to be diagnosed than boys with these disorders who used services (Figure 5, Panel B). Young people from more deprived neighbourhoods who were estimated to have had generalised anxiety disorder or PTSD and to have used health services were less likely to be diagnosed than those from less deprived neighbourhoods with these disorders who used services (Figure 5, Panel C). Those with Black, Asian, Mixed or Other ethnicity who were estimated to have had any of the five disorders and to have used health services were less likely to be diagnosed than their White peers with these disorders who used services (Figure 5, Panel D). These results reflect sociodemographic differences in diagnosis provision within services (after accounting for disorder prevalence and health service use).

## DISCUSSION

In this study of 17-year-olds from South London in 2009-2024, we found that 4.0% of the population had a diagnosis in NHS mental healthcare records during the previous year. Importantly, diagnosis provision increased 1.7-fold over the 16-year period (from 2.6% to 4.4% of the general population), suggesting a substantial increase in the number of young people whose difficulties were recognised in these specialist services. Nevertheless, diagnoses were provided to less than 1 in 16 of those estimated to have had a disorder, and less than 1 in 4 of those estimated to have also used health services for help with their mental health problems. Additionally, diagnosis provision was lower in certain sociodemographic groups; for example, any disorder diagnosis provision was lower in girls than boys and in young people with Black, Asian, Mixed, or Other ethnicity than their White peers, in those estimated to have had a mental health disorder and to have used health services. These findings provide evidence that large diagnosis gaps and biases exist, including within health services, and highlight a vital need for further research and greater investment and equity in young people’s mental healthcare.

Although we endeavoured to use the best available data and methods, limitations should be noted. First, our diagnosis data might be subject to some recording errors. On the one hand, some accurate diagnoses may have been missed if recorded only in unstructured free-text fields of mental health records, as we focused only on diagnoses recorded in structured fields. Natural language processing (NLP) tools have been developed to identify unstructured diagnoses; however, concerns have been raised about the validity of NLP models applied to different samples,^13^ and our pilot testing of the available NLP application for diagnoses found poor precision (appendix p35), so NLP was not used for this study. Furthermore, clinical teams are required to document diagnoses in structured fields before patients’ discharge from the electronic record system, which minimises this limitation. On the other hand, any inaccurate structured diagnoses would have been included in our analyses and treated as if they were accurate, leading us to detect more diagnosis provision than there actually was in Samples 2 and 3, which is unavoidable. Second, differences between the E-Risk Study and the South London population may have introduced bias in results for Samples 2 and 3. To reduce this bias, we weighted calculations according to the sociodemographic characteristics of 17-year-olds in South London. Third, we could not account for changes in disorder and health service use prevalence over time, as this epidemiological data has not been collected. Because disorder prevalence may have increased,^14^ we may have under-estimated disorder prevalence and over-estimated diagnosis provision in more recent years in Samples 2 and 3. These limitations are unlikely to undermine our conclusions.

Our findings are consistent with previous research showing under-diagnosis of mental health disorders in young people. In cohort participants who met criteria for mental health disorders, linkages with health records have found similar rates of diagnosis provision.^15,16^ Additionally, within health services, thorough interviews compared with records suggest large gaps in diagnosis provision.^17^ In line with these findings, the STADIA trial found that patients in English NHS child and adolescent mental health services had high symptom scores and impairment, but low rates of diagnosis provision.^18^ Our research complements these previous findings and provides evidence to counteract recent concerns about over-diagnosis:^19^ although it is possible there is some misdiagnosis, our study and others strongly suggest that there is considerably more under-than over-diagnosis from NHS mental health services for young people.

Our findings have implications for future research, showing that further evaluation of healthcare gaps is required, including research on diagnosis provision in other age groups and regions, as well as research on the provision of evidence-based treatments. Additionally, our findings indicate the importance of investigating the needs of young people who meet criteria for mental health disorders but do not receive diagnoses. It is possible that they have different presentations (e.g., milder difficulties) and different healthcare needs than those who receive diagnoses. However, studies show that people with mild disorders have poor outcomes,^20^ and therefore interventions for this group may be beneficial and cost-effective to treat current symptoms and prevent future deterioration. Moreover, epidemiological research has found that 19% of young people with mental health disorders have serious difficulties,^21^ which is substantially higher than our diagnosis provision rates (6.3%), implying that many severe cases were not diagnosed. Thus, notwithstanding the requirement for further research, our results suggest that NHS mental health service diagnosis provision does not meet need.

Our findings additionally highlight the importance of understanding the effects of diagnosis provision. Qualitative research has found that clinicians, parents, and young people think that accurate diagnosis provision can improve understanding and enable access to effective treatments,^22,23^ in line with the premise of this study. However, clinicians, parents, and young people have also raised concerns about the potential negative effects of labelling, and so some clinicians have reported a reluctance to diagnose.^22,23^ Nevertheless, quantitative research in adult services has found that mental health diagnosis provision is linked with better outcomes.^24^ Further research is therefore needed to investigate this topic and understand how diagnoses can be used most beneficially in young people.

Our research also has implications for clinical practice and policy. Our findings of large diagnosis provision gaps demonstrate the need to reduce barriers to evidence-based healthcare for young people with mental health disorders. Barriers to families seeking help from health services include limited knowledge about mental health and healthcare, stigma, and confidentiality concerns, which are important factors to address.^25^ A key barrier to diagnosis provision within health services is a lack of capacity in specialist mental health teams,^26^ and another possible barrier is insufficient training in referring and specialist services.^26,27^ There is therefore a pressing need to invest in increasing capacity in young people’s mental health services and improving clinician training. Additionally, our findings of diagnosis provision biases, which could reflect inequalities in mental healthcare referrals, access, or assessment,^28,29^ suggest the need to identify and address these challenges and embed equity in the development of youth mental health services. Importantly, reducing barriers and increasing access to equitable diagnosis provision must be accompanied by improved provision of evidence-based treatments. To fully address healthcare gaps, it will also be necessary to develop, evaluate, and implement effective, scalable approaches to early intervention and prevention.^30^

## Data Availability

The South London and Maudsley NHS Foundation Trust (SLaM) Biomedical Research Centre (BRC) Case Register data accessed by the Clinical Record Interactive Search (CRIS) application remain within an NHS firewall and governance is provided by the CRIS Oversight Committee. Subject to these conditions, data are available to researchers upon successful application for access (http://www.maudsleybrc.nihr.ac.uk/facilities/clinical-record-interactive-search-cris). E-Risk Study data are also available to researchers upon successful application for access, according to the E-Risk Study governance processes (https://www.eriskstudy.com/data-access/).

## Contributors

SJL and AD formulated the research questions and designed the analysis plan. AC, TF, KCK, TEM, and MP contributed to the analysis plan. TEM and AC designed the E-Risk Study. TEM, AC, LA, and HLF acquired funding for the E-Risk Study, and supervised data collection and data curation for the E-Risk Study. SJL undertook the data analysis, and AJM checked the data analysis. SJL and AD drafted the manuscript. All authors assisted with interpretation, commented on drafts of the manuscript, and approved the final version.

## Acknowledgements

The SLaM BRC Case Register and CRIS have been developed and are maintained by the National Institute for Health Research (NIHR) Biomedical Research Centre (BRC) at South London and Maudsley NHS Foundation Trust (SLaM) and King’s College London. The E-Risk Study is funded by the UK Medical Research Council (MRC) [G1002190; MR/ X010791/1]; additional support was provided by the USA National Institute of Child Health and Development [HD077482], the Jacobs Foundation, the UK National Society for Prevention of Cruelty to Children (NSPCC), and the UK Economic and Social Research Council (ESRC). SJL has been supported by a Prudence Trust Fellowship and an MRC Clinical Research Training Fellowship [MR/S001492/1]. AD was funded by the MRC [P005918] and the NIHR BRC at SLaM (NIHR203318) and King’s College London. LA has been the Mental Health Leadership Fellow for the ESRC. HLF has been part supported by the ESRC Centre for Society and Mental Health at King’s College London [ES/S012567/1; UKRI861]. JBN has been supported by a Sir Henry Wellcome Postdoctoral Fellowship from the Wellcome Trust [218632/Z/19/Z]. The views expressed are those of the authors and not necessarily those of the NHS, the NIHR, the Department of Health and Social Care, the ESRC, or King’s College London. We are grateful to the E-Risk Study participants for taking part in the research, and to the CRIS and E-Risk Study teams for their dedication, hard work, and insights. We thank Hitesh Shetty for assisting with CRIS data extraction. We also thank Elizaveta Dimitrova and Anya Ruhrberg-Golding for their work piloting natural language processing.

