## Appendix for "Diagnosis provision by young people’s mental health services: a comparison with epidemiological data"

### Appendix: Supplementary Material

### A. Treatments for mental health disorders in young people that are recommended by the UK National Institute of Health and Care Excellence (NICE)

| Disorder | Treatments recommended by NICE | State-funded organisations providing these treatments in South London in 2009-2024 |
|---|---|---|
| Depressive disorder | **NICE guideline 134 – Depression in children and young people: identification and management**.[1] Summary of recommended treatments for 12-18-year-olds:<br>Mild depression: watchful waiting for 2 weeks; if without significant comorbid problems or active suicidal ideas, offer digital cognitive behavioural therapy (CBT), group CBT, group interpersonal psychotherapy (IPT), or group non-directive supportive therapy; if shared decision making based on full assessment (including maturity and developmental level) indicates needs would not be met with previously listed treatments, consider individual CBT or attachment-based family therapy.<br>Moderate to severe depression (or mild depression that continues after 2-3 months of previously listed psychological therapy): offer individual CBT for at least 3 months; with or without fluoxetine medication; if shared decision making based on full assessment (including maturity and developmental level) indicates needs would not be met with individual CBT, consider IPT for adolescents (IPT-A), family therapy (attachment-based or systemic), brief psychosocial intervention, or psychodynamic psychotherapy; with or without fluoxetine medication.<br>Depression unresponsive to treatment / recurrent depression / psychotic depression: an alternative psychological therapy not previously tried; with or without fluoxetine, sertraline, or citalopram medication, and augmentation with an antipsychotic medication.<br>All: if an antidepressant medication is prescribed, this should only be following assessment and diagnosis by a child and adolescent psychiatrist. | South London and Maudsley NHS Foundation Trust, child and adolescent mental health services (range of teams, from those offering low intensity interventions to those providing the most complex care)<br><br>Mental Health Support Teams in schools and colleges, delivered by South London and Maudsley NHS Foundation Trust (use the electronic health record system) in Croydon, Lambeth, and Lewisham, but are separate in Southwark (these teams deliver low intensity interventions, such as guided self-help based on CBT) |
| Generalised anxiety disorder | **There is no NICE guideline for generalised anxiety disorder in young people**.<br>**NICE guideline 113 – Generalised anxiety disorder and panic disorder in adults: management**.[2] Summary of recommended treatments for generalised anxiety disorder (GAD) in adults:<br>Provide education and active monitoring; offer individual non-facilitated self-help, guided self-help based on CBT, or psychoeducational groups, that are based on CBT principles with structured materials for a period of 6 weeks.<br>GAD with marked functional impairment or that has not improved after previously listed interventions: offer an individual high-intensity psychological intervention (CBT or applied relaxation) or drug treatment (consider offering sertraline first).<br>Inadequate response: Offer an alternative high-intensity psychological intervention or drug treatment, or combination.<br>**Americal Academy of Child and Adolescent Psychiatry Clinical Practice Guideline for the assessment and treatment of children and adolescents with anxiety disorders**.[3] Summary of recommended treatments for GAD in 6-18-year-olds:<br>Offer CBT.<br>Offer a selective serotonin reuptake inhibitor (SSRI) medication.<br>Combination CBT and SSRI could be offered preferentially over CBT alone or an SSRI alone.<br>Serotonin norepinephrine reuptake inhibitor medication could be offered. | South London and Maudsley NHS Foundation Trust, child and adolescent mental health services (range of teams, from those offering low intensity interventions to those providing the most complex care)<br><br>Mental Health Support Teams in schools and colleges, delivered by South London and Maudsley NHS Foundation Trust (use the electronic health record system) in Croydon, Lambeth, and Lewisham, but are separate in Southwark (these teams deliver low intensity interventions, such as guided self-help based on CBT) |
| Post-traumatic stress disorder | **NICE guideline 116 – Post-traumatic stress disorder**.[4] Summary of recommended treatments for 7-17-year-olds:<br>3+ months after trauma: offer individual trauma-focused CBT; if they do not respond or engage with individual trauma-focused CBT, consider eye movement desensitisation and reprocessing.<br>1-3 months after trauma: consider individual trauma-focused CBT. | South London and Maudsley NHS Foundation Trust, child and adolescent mental health services<br><br>Other specialist services deliver only a small proportion of PTSD care, such as sexual assault referral centres |
| Conduct disorder | **NICE clinical guideline 158 – Antisocial behaviour and conduct disorders in children and young people: recognition and management**.[5] Summary of recommended treatments for 11-17-year-olds:<br>Offer multimodal intervention, e.g., multisystemic therapy.<br>Young people with conduct disorder who have problems with explosive anger and severe emotional dysregulation and who have not responded to psychosocial interventions: consider risperidone medication for the short-term management of severely aggressive behaviour. | South London and Maudsley NHS Foundation Trust, child and adolescent mental health services<br><br>South London local authorities |

Table continued.

| Disorder | Treatments recommended by NICE | State-funded organisations providing these treatments in South London in 2009-2024 |
|---|---|---|
| Attention-deficit/hyperactivity disorder (ADHD) | **NICE guideline 87 – Attention deficit hyperactivity disorder: diagnosis and management**.[6] Summary of recommended treatments for 5+-year-olds: Give information about ADHD, advice on parenting strategies, and liaise with school; if also have symptoms of conduct disorder, offer parent training programme, or if difficulties attending group/needs too complex, consider individual parent training programme; if persistent significant impairment after environmental modifications have been implemented and reviewed, offer medication (offer methylphenidate as first line); for young people with ADHD who have benefited from medication but whose symptoms are still causing a significant impairment, consider CBT for ADHD. | South London and Maudsley NHS Foundation Trust, child and adolescent mental health services<br><br>South London community paediatric services deliver a small proportion of ADHD care |

The table outlines treatments that are recommended by NICE for young people with the five mental health disorders that we focused on in this study, and the state-funded organisations that provided these treatments in South London during our study time period (2009-2024). NICE recommendations are developed based on evidence of clinical- and cost-effectiveness. Of note, the skills and ongoing supervision required for these treatments are largely only available in specialist mental health services, and these state-funded services are provided by South London and Maudsley NHS Foundation Trust (SLaM) in South London. In contrast, schools, general practices, community hubs, and charities have mostly provided other interventions (e.g. supportive counselling) which may be appropriate for subthreshold symptoms, but are not recommended for the treatment of mental health disorders. Mental Health Support Teams (MHSTs) in schools began in South London from 2019, to provide low intensity evidence-based interventions (e.g. guided self-help based on CBT) to those with mild problems. MHSTs are provided by SLaM in Croydon, Lambeth, and Lewisham, but are separate in Southwark.

### B. Definitions of mental health disorders used in this study

| Disorder | Data source for mental health diagnosis provision: South London and Maudsley NHS Foundation Trust clinical records | Data source for mental health disorders and health service use: E-Risk Study |
|---|---|---|
| Depressive disorder | ICD-10 F31.3-31.5 Bipolar affective disorder, current episode depression;<br>ICD-10 F32 Depressive episode;<br>ICD-10 F33 Recurrent depressive disorder;<br>ICD-10 F92.0 Depressive conduct disorder | DSM-IV Major depressive episode |
| Generalised anxiety disorder | ICD-10 F41.1 Generalized anxiety disorder | DSM-IV Generalized anxiety disorder |
| Other anxiety disorder | ICD-10 F40 Phobic anxiety disorders;<br>ICD-10 F41 Other anxiety disorders (excluding F41.1 Generalized anxiety disorder);<br>ICD-10 F93.0 Separation anxiety disorder of childhood;<br>ICD-10 F93.1 Phobic anxiety disorder of childhood;<br>ICD-10 F93.2 Social anxiety disorder of childhood | - |
| Obsessive-compulsive disorder | ICD-10 F42 Obsessive-compulsive disorder | - |
| Post-traumatic stress disorder | ICD-10 F43.1 Post-traumatic stress disorder | DSM-5 Post-traumatic stress disorder |
| Other stress-related disorder | ICD-10 F43 Reaction to severe stress, and adjustment disorders (excluding F43.1 Post-traumatic stress disorder) | - |
| Dissociative or somatoform disorder | ICD-10 F44 Dissociative [conversion] disorders;<br>ICD-10 F45 Somatoform disorders | - |
| Eating disorder | ICD-10 F50 Eating disorders | - |
| Conduct disorder | ICD-10 F90.1 Hyperkinetic conduct disorder;<br>ICD-10 F91 Conduct disorders;<br>ICD-10 F92 Mixed disorders of conduct and emotions | DSM-IV Conduct disorder |
| Substance use disorder | ICD-10 F10-19 Mental and behavioural disorders due to psychoactive substance use | - |
| Intellectual disability | ICD-10 F70-79 Mental retardation | - |
| Autistic spectrum disorder | ICD-10 F84 Pervasive developmental disorders | - |
| Attention-deficit/hyperactivity disorder | ICD-10 F90 Hyperkinetic disorders | DSM-5 Attention-deficit/hyperactivity disorder |
| Psychotic disorder | ICD-10 F20-29 Schizophrenia, schizotypal and delusional disorders | - |
| Mania or bipolar disorder | ICD-10 F30 Manic episode;<br>ICD-10 F31 Bipolar affective disorder (excluding F31.3-31.5 Bipolar affective disorder, current episode depression) | - |

The table outlines the definitions of mental health disorders used in this study. For diagnosis provision from South London and Maudsley NHS Foundation Trust (SLaM) electronic health records, disorders were diagnosed and recorded in structured fields by clinicians according to the International Statistical Classification of Diseases and Related Health Problems 10th Revision (ICD-10).[7] For estimated disorder prevalence in the population based on E-Risk Study data, disorders were systematically assessed by researchers according to Diagnostic and Statistical Manual of Mental Disorders (DSM-IV or -5) criteria for depressive disorder, generalised anxiety disorder, post-traumatic stress disorder, conduct disorder, and attention-deficit/hyperactivity disorder.[8–13]

To prevent under-estimation of diagnosis provision in Samples 2 and 3 resulting from differences in diagnostic systems used by SLaM and the E-Risk Study, we included a wide range of SLaM diagnosis ICD-10 codes for each disorder, including 'unspecified' categories, which have lower thresholds than the DSM criteria used in the E-Risk Study.

### C. Data source for mental health diagnosis provision: South London and Maudsley NHS Foundation Trust, additional information

We obtained data about mental health diagnosis provision from South London and Maudsley NHS Foundation Trust, which was extracted via the Maudsley Biomedical Research Centre Case Register using the Clinical Record Interactive Search application, as outlined in the manuscript. This organisation is the sole provider of state-funded universal specialist mental healthcare for the population of four South London boroughs (Croydon, Lambeth, Lewisham, and Southwark; referred to as 'South London' throughout). This data source has been described in detail in previous papers.[14–16] It has been approved for secondary analysis by the South Central - Oxford C Research Ethics Committee (reference 23/SC/0257).

We selected data on young people from South London who were aged 17 (i.e., 17≤age<18) years on 30th June each year in 2009-2024 and had an active mental health diagnosis during the previous year. We ensured they were from South London (Croydon, Lambeth, Lewisham, and Southwark) by restricting the dataset to young people who were registered with South London general practitioners (GPs; criterion that determines NHS mental healthcare provider) at the time of diagnosis (or within one month); or if GP details were missing, who lived in South London within at the time of diagnosis (or within one month). In this way, we avoided over-estimation of diagnosis provision attributable to out-of-area patients, such as those attending national and specialist services that are delivered by this provider. We chose age 17 years to capture those who received care from child and adolescent mental health services and to best fit with data for estimated prevalences of disorders and health service use (described in the manuscript and below). We chose 30th June annually to align with data for population characteristics (described in the manuscript and below). We chose 2009 as the start of our study period because 2008 is the first year that electronic health records were used by all teams in South London and Maudsley NHS Foundation Trust. We defined active diagnosis as starting on the date recorded by clinicians and ending either on the date of a subsequent mental health diagnosis that did not contain the disorder (including a diagnosis of no mental health disorder) or on the date of their discharge from mental health services, whichever was first.

We extracted these patients' diagnosis data from structured fields, coded by clinicians for each patient, according to the International Statistical Classification of Diseases and Related Health Problems 10th Revision (ICD-10),[7] which must be completed before patients can be discharged. We included diagnoses of a wide range of mental health disorders (as detailed above in appendix p4). We also extracted patients' sociodemographic data from structured fields, including gender, ethnicity, and home neighbourhood (lower-layer super output area [LSOA], comprising approximately 1,500 residents) at diagnosis. To determine patients' neighbourhood deprivation level, we identified their home neighbourhood's Index of Multiple Deprivation (IMD), the UK Government's measure of deprivation in English neighbourhoods, categorised in national quintiles. For patients with diagnoses on or before 30th June 2017, we defined their home neighbourhood based on LSOA 2011 (built from 2011 census areas), and mapped this to IMD 2015.[17] For patients with diagnoses between 1st July 2017 and 30th June 2020, we defined their home neighbourhood based on LSOA 2011, and mapped this to IMD 2019.[18] For patients with diagnoses on or after 1st July 2020, we defined their home neighbourhood based on LSOA 2021 (built from 2021 census areas), and mapped this to IMD 2025.[19] This method was chosen to align with available population data (described below in appendix p6).

### D. Data source for population characteristics: UK Office for National Statistics, additional information

We obtained general population data from the UK Office for National Statistics, which estimates population size as of 30th June each year, based on the previous census and subsequent administrative data.[20] We sought the number of 17-year-olds who lived in South London (Croydon, Lambeth, Lewisham, and Southwark) in 2009-2024, including by sex (as a proxy for gender) and neighbourhood (lower-layer super output area [LSOA]). For 2009-2020, we defined South London and LSOAs according to 2011 census areas; and for 2021-2024, we defined South London and LSOAs according to 2021 census areas, based on available data. Neighbourhood deprivation in this population was determined by identifying Index of Multiple Deprivation national quintiles corresponding to these neighbourhoods. For 2009-2017, we mapped LSOA 2011 to IMD 2015.[17] For 2018-2020, we mapped LSOA 2011 to IMD 2019.[18] For 2021-2024, we mapped LSOA 2021 to IMD 2025.[19] Ethnicity of the population was estimated by applying ethnicity ratios found in censuses. For 2009-2015, we used ethnicity ratios from the 2011 census, based on data about 16-17 year-olds in South London.[21] For 2016-2024, we used ethnicity ratios from the 2021 census, based on data about 17 year-olds in South London.[22]

### E. Data source for mental health disorders and health service use: epidemiological study review and selection

#### Prespecified criteria

We sought to derive estimated prevalence rates of mental health disorders and health service use from existing epidemiological research. We reviewed available epidemiological studies against criteria that we prespecified to maximise accuracy, including essential criteria that we decided must be met and desirable criteria from which we planned to select the most suitable study:

*Essential criteria*

Sample characteristics:

- Age at recruitment ≤18 years – given our focus on young people, and that our data source for diagnosis provision captures most state-funded healthcare provision leading to evidence-based treatments for local <18-year-olds with mental health disorders.
- South London or nationwide coverage – so that data are comparable with our data source for diagnosis provision, or similar to and can be weighted to match the sociodemographic characteristics of young people from South London.
- Population-based study – including recruited from population registers or common settings e.g. schools, to reflect the general population.

Mental health assessment:

- Mental health disorder assessment age ≤18 years – assessing whether young people met established diagnostic criteria, i.e. ICD or DSM (rather than symptom scores).
- Mental health disorder assessment since 2009 – assessing participants since the start of our study period, to provide contemporaneous prevalence estimates that overlap with the timing of our data source for diagnosis provision.
- Mental health disorder assessment with interview – assessment using structured interviews based on diagnostic criteria. These interviews allow explanation and confirmation of understanding and therefore are less likely to result in misinterpretation than questionnaires. These interviews are also feasible for large-scale epidemiological studies, whereas clinical assessments are not. Most previous research has found that these interviews have moderate to substantial agreement with blinded clinical reappraisal interviews and provide fairly similar prevalence estimates.[23–25] Some studies that have shown poorer agreement have found lower prevalence estimates from structured interviews compared with clinical interviews, i.e. this method might produce more conservative prevalence estimates.[26,27]
- Health service use for mental health, assessment at same time – given our interest in considering diagnosis provision in those who use health services.

*Desirable criteria*

Sample characteristics:

- Sample size large – large samples are more likely to provide reliable estimates of the prevalence of mental health disorders and health service use, including in subgroups based on sociodemographic characteristics. We considered a sample size of over 2,000 would be desirable.
- Response/retention rate high – the majority of the sampling frame invited to participate should have taken part to minimise non-responder bias and therefore maximise validity. Research shows that non-responders have higher rates of mental health disorders than responders.[28]
- Sociodemographic representativeness high – sociodemographic characteristics similar to the general population, to maximise validity and generalisability of findings.

Mental health assessment:

- Disorders assessed by interview wide-ranging – covering multiple common internalising and externalising/neurodevelopmental conditions, according to established diagnostic criteria, i.e. ICD or DSM.
- Service use for mental health problems comprehensively assessed by interview – covering a range of different healthcare clinicians and settings.
- Informant young person – young person or multi-informant methods preferred to parent-only reports. Research shows poor concordance between young person and parent reports in assessments of mental health disorders,

particularly for internalising disorders,[29] with parents under-reporting problems perhaps because they are not fully aware of the young person's internal experiences.

- Reporting term 12 months – 12 months is preferred to point prevalence, because it provides more stable estimates.[30]
- Year(s) assessed over 2009-2024 – ideally assessments would have been repeated in multiple years during our study period, to enable us to account for changes in prevalence of mental health disorders and services use over time. If multiple assessments are not available, we will consider one estimate within our study period and acknowledge this limitation.

### Review

We reviewed existing epidemiological studies, including systematically reviewing the Catalogue of Mental Health Measures for studies with age at recruitment from birth to 18 years.[31] We assessed whether each study met our essential criteria, and if so we considered our desirable criteria. The results of this review are outlined in the table below. Panel 1 presents the studies that met essential criteria. Panel 2 presents the studies that did not meet our essential criteria. Our selection considerations are described after the table.

| Study | Criteria |
|---|---|
| **1. Studies that met our essential criteria** | |
| **Environmental Risk (E-Risk) Longitudinal Twin Study**[12,32] | *Essential criteria*<br>Sample characteristics:<br>• Age at recruitment ≤18 years: Yes, 5 years<br>• South London or nationwide coverage: Yes, nationwide England and Wales<br>• Population-based study: Yes, recruited from a birth register of twins born in England and Wales in 1994 and 1995<br>Mental health assessment:<br>• Mental health disorder assessment age ≤18 years: Yes, 18 years<br>• Mental health disorder assessment since 2009: Yes, 2012-2013<br>• Mental health disorder assessment with interview: Yes<br>• Health service use for mental health, assessment at same time: Yes<br><br>*Desirable criteria*<br>Sample characteristics:<br>• Size: 2,066<br>• Response/retention: 93% of original sample were followed up at age 18, 93% of those targeted were included in original sample, i.e., 86% of those originally targeted were followed up at age 18<br>• Sociodemographic representativeness: families were recruited to represent the UK population with newborns in the 1990s. The sample represents the full range of socioeconomic conditions in Great Britain; the families' distribution is reflected on a neighbourhood-level socioeconomic index (ACORN) that very closely matches the national distribution. The sample is comprised of twins, but previous research has found no substantial difference in rates of emotional and behavioural psychopathology between twins and singletons,[33–35] supporting the generalisability of findings from twin studies to the general population.<br>Mental health assessment:<br>• Disorders: depressive disorder, generalised anxiety disorder, post-traumatic stress disorder, conduct disorder, attention-deficit/hyperactivity disorder, assessed during face-to-face interviews using the Diagnostic Interview Schedule for DSM-IV or adapted structured interviews to ascertain DSM-5 diagnostic criteria.<br>• Service use: questions enquiring whether participants had seen a range of health professionals (including medical doctor, GP; a psychologist, counsellor, or therapist; or a psychiatrist) for mental health problems asked as part of the interview<br>• Informant: Young person<br>• Reporting term: 12 months<br>• Assessment year(s): 2012-20213 (single assessment) |
| **IMAGEN**[36,37] | *Essential criteria*<br>Sample characteristics:<br>• Age at recruitment ≤18 years: Yes, 14 years<br>• South London or nationwide coverage: Yes, London (in addition to Nottingham and other European cities: Dublin, Paris, Berlin, Hamburg, Mannheim and Dresden)<br>• Population-based study: Yes, recruited from high schools<br>Mental health assessment:<br>• Mental health disorder assessment age ≤18 years: Yes, 14 years and follow up at 16 years<br>• Mental health disorder assessment since 2009: Yes, 2008-2010 and follow up in 2011-2012<br>• Mental health disorder assessment with interview: Yes<br>• Health service use for mental health, assessment at same time: Yes |

Table continued

| Study | Criteria |
|---|---|
| **IMAGEN[36,37] continued** | *Desirable criteria*<br>Sample characteristics:<br>• Size: ~250 in London (~2,000 over 8 European sites) at baseline<br>• Response/retention: Not stated<br>• Sociodemographic representativeness: "To obtain a diverse sample in terms of SES, emotional and cognitive development, private, state-funded and special units are equally targeted. To maximize ethnic (Caucasian) homogeneity, at each study centre recruitment focuses on geographical areas with minimal ethnic diversity."<br>Mental health assessment:<br>• Disorders: separation anxiety disorder, specific phobias, social phobia, panic disorder, agoraphobia, post-traumatic stress disorder, obsessive-compulsive disorder, depression, bipolar disorder, attentional and hyperkinetic disorders, conduct disorders, eating disorders, tic disorders (young person and parent) and autism spectrum disorder (parent only), assessed using the Development and Wellbeing Assessment (DAWBA)<br>• Service use: questions enquiring whether participants had seen a range of health professionals (including GP, family doctor, health visitor; counsellor, psychologist, psychiatrist) for mental health problems asked in the DAWBA (parent only)<br>• Informant: young person and parent for mental health disorders, parent only for service use<br>• Reporting term: current for mental health disorders, and not stated for service use<br>• Assessment year(s): 2008-2010 and follow up in 2011-2012 |
| **Mental Health of Children and Young People in Great Britain (MHCYP)[38]** | *Essential criteria*<br>Sample characteristics:<br>• Age at recruitment ≤18 years: Yes, 2-19 years (2017)<br>• South London or nationwide coverage: Yes, nationwide England<br>• Population-based study: Yes, stratified multistage random probability sample of children was drawn from NHS patient register<br>Mental health assessment:<br>• Mental health disorder assessment age ≤18 years: Yes, 2-19 years<br>• Mental health disorder assessment since 2009: Yes, 2017 (all other surveys completed before 2009, or have only considered problems/symptoms rather than disorder criteria)<br>• Mental health disorder assessment with interview: Yes, 2017<br>• Health service use for mental health, assessment at same time: Yes<br><br>*Desirable criteria*<br>Sample characteristics:<br>• Size: 9,117<br>• Response/retention: 51% response rate<br>• Sociodemographic representativeness: the sampling frame was selected to be representative of children and young people in England, and the data analysis was weighted so that results are representative (accounting for those who did not take part in the survey)<br>Mental health assessment:<br>• Disorders: separation anxiety disorder, specific phobia, social phobia, panic disorder, agoraphobia, post-traumatic stress disorder, obsessive-compulsive disorder, body dysmorphic disorder, generalised anxiety disorder, depressive disorder, disruptive mood dysregulation disorder, hyperactivity disorder, behavioural (conduct) disorder, pervasive developmental disorders, eating disorders, tics, bipolar affective disorder, according to ICD-10 and DSM-IV or DSM-5 assessed using the Development and Wellbeing Assessment (DAWBA)<br>• Service use: questions enquiring whether participants had seen a range of health professionals (including primary care specialist; and mental health specialist) for mental health problems asked during the interview<br>• Informant: young person (11-19-year-olds) and parent (2-19-year-olds)<br>• Reporting term: current for mental health disorders, and past year for service use<br>• Assessment year(s): 2017 |
| **Resilience, Ethnicity & Adolescent Mental Health (REACH)[39]** | *Essential criteria*<br>Sample characteristics:<br>• Age at recruitment ≤18 years: Yes, 11-14 years<br>• South London or nationwide coverage: Yes, South London, Southwark and Lambeth<br>• Population-based study: Yes, recruited from secondary schools<br>Mental health assessment:<br>• Mental health disorder assessment age ≤18 years: Yes, 11-14 (part 2 participants only), 12-15 years follow up (part 2 participants only)<br>• Mental health disorder assessment since 2009: Yes, 2016-2017, 2017-2018 follow up<br>• Mental health disorder assessment with interview: Yes<br>• Health service use for mental health, assessment at same time: Yes |

Table continued

| Study | Criteria |
|---|---|
| **Resilience, Ethnicity & Adolescent Mental Health (REACH)[39] continued** | *Desirable criteria*<br>Sample characteristics:<br>• Size: 803 (part 2 interviews)<br>• Response/retention: 88% (4,353/4,945) response rate for part 1 questionnaires; of these, consent for part 2 interviews was obtained from 21% (1,060/4,353); of these, 803 completed part 2 (/1,060=76%, although it is not clear if all were asked); of these, 74% (552/803) took part in 1-year follow-up<br>• Sociodemographic representativeness: Schools were selected to be representative of the 38 mainstream secondary schools within the two boroughs based on (i) the proportion of students eligible for free school meals (a marker of household socioeconomic disadvantage) and (ii) the proportion of students in minority ethnic groups. The REACH cohorts (part 1) are highly representative of the target population. However, less information is available on the representativeness of the part 2 interview sub-sample.<br>Mental health assessment:<br>• Disorders: post-traumatic stress disorder, anxiety, attention-deficit/hyperactivity disorder, conduct disorder, assessed using the Development and Wellbeing Assessment (DAWBA)<br>• Service use: help seeking was assessed during the part 2 interview<br>• Informant: young person<br>• Reporting term: current for mental health disorders, and not reported for service use<br>• Assessment year(s): 2016-2017, 2017-2018 follow up |
| **2. Studies that do not meet our essential criteria** | |
| **ADVANCE: Armed Services Trauma Rehabilitation Outcome Study** | • Age at recruitment ≤18 years: No, 18-50 years (at sampling) |
| **Aetiology and Ethnicity in Schizophrenia and Other Psychoses (AESOP-10)** | • Age at recruitment ≤18 years: No, 16-65 years, primarily focused on adults |
| **The Airwave Health Monitoring Study** | • Age at recruitment ≤18 years: No, 16-80 years, primarily focused on adults |
| **Avon Longitudinal Study of Parents & Children (ALSPAC)** | • Age at recruitment ≤18 years: Yes, birth<br>• South London or nationwide coverage: No, Bristol and District Health Authority |
| **Adult Psychiatric Morbidity Survey (APMS)** | • Age at recruitment ≤18 years: No, 16+ years, primarily focused on adults |
| **Surveys of Psychiatric Morbidity** | • Age at recruitment ≤18 years: No, 16+ years, primarily focused on adults |
| **British Autism Study of Infant Siblings (BASIS)** | • Age at recruitment ≤18 years: Yes, birth (from prenatal to early infancy)<br>• South London or nationwide coverage: Yes, nationwide<br>• Population-based study: No, Phase 1–2=children at elevated likelihood for ASD, Phase 3=children at elevated likelihood for ASD, ADHD, or both |
| **1970 British Cohort Study (BCS70)** | • Age at recruitment ≤18 years: Yes, birth<br>• South London or nationwide coverage: Yes, nationwide<br>• Population-based study: Yes, birth cohort<br>• Mental health disorder assessment age ≤18 years: No, problems/symptoms only, at age 5, 10, 16 years<br>• Mental health disorder assessment since 2009: No, problems/symptoms most recently 1986 (16 years old) |
| **Born in Bradford (BiB)** | • Age at recruitment ≤18 years: Yes, birth<br>• South London or nationwide coverage: No, Bradford |
| **Born in Bradford's Better Start (BiBBS)** | • Age at recruitment ≤18 years: Yes, birth<br>• South London or nationwide coverage: No, Bradford |
| **COVID Social Mobility & Opportunities (COSMO) Study** | • Age at recruitment ≤18 years: Yes, 16-17 years<br>• South London or nationwide coverage: Yes, nationwide<br>• Population-based study: Yes, students who were in Year 11<br>• Mental health disorder assessment age ≤18 years: No, problems/symptoms only, at age 15-16 and 17-18 years<br>• Mental health disorder assessment since 2009: No, problems/symptoms only, in 2021-2022 and 2022-2023<br>• Mental health disorder assessment with interview: No, questionnaires completed online |
| **Clinical Record Interactive Search (CRIS)** | • Age at recruitment ≤18 years: Yes, varied<br>• South London or nationwide coverage: Yes, South London<br>• Population-based study: No, individuals who have accessed the services of South London and Maudsley (SLaM) NHS Foundation Trust |

Table continued

| Study | Criteria |
|---|---|
| **The Cambridge Study in Delinquent Development (CSDD)** | • Age at recruitment ≤18 years: Yes, 8-9 years<br>• South London or nationwide coverage: Yes, South London<br>• Population-based study: Yes, males born in 1952–53 from a working-class area of South London<br>• Mental health disorder assessment age ≤18 years: Possibly problems/symptoms and doctor diagnoses only (detail not available), at age 8, 10, 12, 14, 16, 18 years<br>• Mental health disorder assessment since 2009: No, most recently 1971-1972 (18 years old) |
| **Determinants of Adolescent Social Wellbeing & Health (DASH)** | • Age at recruitment ≤18 years: Yes, 11-13 years<br>• South London or nationwide coverage: Yes, London<br>• Population-based study: Yes, pupils from 51 schools across 10 inner London boroughs<br>• Mental health disorder assessment age ≤18 years: No, problems/symptoms only, at age 11-13, 14-16 years<br>• Mental health disorder assessment since 2009: No, problems/symptoms only, most recently 2005-2006 (14-16 years old) |
| **Education and Child Health Insights from Linked Data (ECHILD)** | • Age at recruitment ≤18 years: Yes, 0-38 years<br>• South London or nationwide coverage: Yes, nationwide<br>• Population-based study: Yes, all children born between 1 September 1984 and 31 August 2022, who have records on the Hospital Episode Statistics (HES) and National Pupil Database (NPD)<br>• Mental health disorder assessment age ≤18 years: Yes, varied<br>• Mental health disorder assessment since 2009: Yes, 2007-2020, 2016<br>• Mental health disorder assessment with interview: No, administrative health records only |
| **Early Prediction of Adolescent Depression (EPAD)** | • Age at recruitment ≤18 years: Yes, 9-17 years<br>• South London or nationwide coverage: No, South Wales |
| **European Quality of Life Survey (EQLS)** | • Age at recruitment ≤18 years: No, 18+ years, primarily focused on adults |
| **European Social Survey (ESS)** | • Age at recruitment ≤18 years: No, 15+ years, primarily focused on adults |
| **The Edinburgh Study of Youth Transitions and Crime (ESYTC)** | • Age at recruitment ≤18 years: Yes, 12 years<br>• South London or nationwide coverage: No, Edinburgh |
| **European Working Conditions Survey (EWCS)** | • Age at recruitment ≤18 years: No, 15+ years, primarily focused on adults |
| **Gemini** | • Age at recruitment ≤18 years: Yes, 8 months<br>• South London or nationwide coverage: Yes, nationwide<br>• Population-based study: Yes, birth cohort (twins)<br>• Mental health disorder assessment age ≤18 years: No, problems/symptoms only, at age 5 and 11-12 years<br>• Mental health disorder assessment since 2009: No, problems/symptoms only, in 2012-2013 and 2019 |
| **Genetic Links to Anxiety and Depression (GLAD) Study** | • Age at recruitment ≤18 years: No, 16+ years, primarily focused on adults |
| **Generation Scotland: Scottish Family Health Study (GS:SFHS)** | • Age at recruitment ≤18 years: No, 18-98 years, primarily focused on adults |
| **Growing Up in Scotland: Birth Cohort 1 and Birth Cohort 2** | • Age at recruitment ≤18 years: Yes, birth<br>• South London or nationwide coverage: No, Scotland |
| **Growing Up in Scotland: Child Cohort** | • Age at recruitment ≤18 years: Yes, 3 years<br>• South London or nationwide coverage: No, Scotland |
| **Hertfordshire Cohort Study (HCS)** | • Age at recruitment ≤18 years: Yes, birth<br>• South London or nationwide coverage: No, Hertfordshire |
| **Intellectual Disability and Mental Health: Assessing the Genomic Impact on Neurodevelopment (IMAGINE ID)** | • Age at recruitment ≤18 years: Yes, 4-19 years<br>• South London or nationwide coverage: Yes, nationwide<br>• Population-based study: No, individuals with intellectual disability of identified genetic aetiology |

Table continued

| Study | Criteria |
|---|---|
| **Health and Wellbeing Cohort Study** | • Age at recruitment ≤18 years: No, 18+ years, primarily focused on adults |
| **Lothian Birth Cohort 1921 (LBC1921)** | • Age at recruitment ≤18 years: Yes, 10-11 years<br>• South London or nationwide coverage: No, Edinburgh and the Lothians, Scotland |
| **Lothian Birth Cohort 1936 (LBC1936)** | • Age at recruitment ≤18 years: Yes, 10-11 years<br>• South London or nationwide coverage: No, Edinburgh and the Lothians, Scotland |
| **Longitudinal Outcomes of Gender Identity in Children (LOGIC)** | • Age at recruitment ≤18 years: Yes, 3-14 years<br>• South London or nationwide coverage: Yes, nationwide<br>• Population-based study: No, children and young people referred to the Gender Identity Development Service |
| **Millennium Cohort Study (MCS)** | • Age at recruitment ≤18 years: Yes, birth<br>• South London or nationwide coverage: Yes, nationwide<br>• Population-based study: Yes, birth cohort<br>• Mental health disorder assessment age ≤18 years: No, problems/symptoms or doctor diagnosis only, at age 5, 7, 11, 14, 17 years<br>• Mental health disorder assessment since 2009: No, problems/symptoms or doctor diagnosis only in 2012-2013 (11 years old), 2015-2016 (14 years old), 2018-2019 (17 years old) |
| **The National Surveys of Sexual Attitudes and Lifestyles (NATSAL)** | • Age at recruitment ≤18 years: No, 16+ years, primarily focused on adults |
| **1958 National Child Development Study (NCDS)** | • Age at recruitment ≤18 years: Yes, birth<br>• South London or nationwide coverage: Yes, nationwide<br>• Population-based study: Yes, birth cohort<br>• Mental health disorder assessment age ≤18 years: No, problems/symptoms only, at age 7, 11, 16 years<br>• Mental health disorder assessment since 2009: No, problems/symptoms only, most recently in 1974 (16 years old) |
| **Next Steps** | • Age at recruitment ≤18 years: Yes, 3-14 years<br>• South London or nationwide coverage: Yes, nationwide<br>• Population-based study: Yes, representative of young people in Year 9 (or equivalent) in England<br>• Mental health disorder assessment age ≤18 years: No, problems/symptoms only, at age 14, 15, 16, 17 years<br>• Mental health disorder assessment since 2009: No, problems/symptoms only, most recently in 2007 (17 years old) |
| **MRC National Survey of Health and Development (NSHD)** | • Age at recruitment ≤18 years: Yes, birth<br>• South London or nationwide coverage: Yes, nationwide<br>• Population-based study: Yes, birth cohort<br>• Mental health disorder assessment age ≤18 years: No, problems/symptoms only, at age 7-18<br>• Mental health disorder assessment since 2009: No, problems/symptoms only, most recently in 1964 (18 years old) |
| **The ROOTS Study** | • Age at recruitment ≤18 years: Yes, 14 years<br>• South London or nationwide coverage: No, Cambridgeshire |
| **The Study of Cognition, Adolescents and Mobile Phones (SCAMP)** | • Age at recruitment ≤18 years: Yes, 11-12 years<br>• South London or nationwide coverage: Yes, Greater London<br>• Population-based study: Yes, secondary school pupils from eligible schools<br>• Mental health disorder assessment age ≤18 years: No, problems/symptoms and self-reports of disorder only, at age 11-12, 13-15, 16-18 years<br>• Mental health disorder assessment since 2009: No, problems/symptoms and self-reports of disorder only, in 2014-2016 (11-12 years old), 2016-2018 (13-15 years old), 2020-2021 (16-18 years old) |
| **South East London Community Health (SELCoH) Study** | • Age at recruitment ≤18 years: No, 16+ years, primarily focused on adults |
| **Social and Economic Predictors of the Severe Mental Disorders (SEP-MD) Study** | • Age at recruitment ≤18 years: Yes, Varied<br>• South London or nationwide coverage: Yes, South London<br>• Population-based study: No, SLaM service users with a diagnosis of severe mental illness |
| **Twins Early Development Study (TEDS)** | • Age at recruitment ≤18 years: Yes, birth<br>• South London or nationwide coverage: Yes, nationwide<br>• Population-based study: Yes, birth cohort<br>• Mental health disorder assessment age ≤18 years: No, problems/symptoms only, at age 2, 3, 4, 7, 9, 12, 14, 16, 18 years<br>• Mental health disorder assessment since 2009: No, problems/symptoms only, in 2007-2011 (age 14 years) 2010-2013 (16 years old), 2012-2014 (18 years old) |

Table continued

| Study | Criteria |
|---|---|
| **TwinsUK: The UK Adult Twin Registry** | • Age at recruitment ≤18 years: No, 18+ years, primarily focused on adults |
| **Understanding Society, the UK Household Longitudinal Survey & British Household Panel Survey (UKHLS)** | • Age at recruitment ≤18 years: Yes, varied including 10-15 years<br>• South London or nationwide coverage: Yes, nationwide<br>• Population-based study: Yes, representative probability sample of households<br>• Mental health disorder assessment age ≤18 years: No, problems/symptoms only<br>• Mental health disorder assessment since 2009: No, problems/symptoms only |
| **Wirral Child Health & Development Study (WCHADS)** | • Age at recruitment ≤18 years: Yes, 20 weeks gestation<br>• South London or nationwide coverage: No, Wirral, Merseyside |
| **West of Scotland Twenty-07** | • Age at recruitment ≤18 years: Yes (youngest cohort), 15, 35, 55 years old<br>• South London or nationwide coverage: No, Central Clydeside Conurbation, West of Scotland |

### Selection

We considered the findings of our review and judged that the study that met all essential criteria and best met desirable criteria was the E-Risk Study. This study has a large sample size (>2,000), high retention/response rate (86% of those originally targeted were still taking part when mental health assessments took place at age 18 years), and is representative of the general population. Additionally, the study assessed multiple common internalising and externalising/neurodevelopmental mental health disorders and service use, by validated interviews with young people. Furthermore, the reporting term was 12 months, which provides more stable estimates than point prevalence rates. Although the study assessed participants for mental health disorders at one time point only in 2012-2013, there was no more suitable study which undertook repeated assessments that spanned our study period. Further information about the E-Risk Study is provided below in appendix p14.

We selected the E-Risk Study over the IMAGEN Study, because the IMAGEN Study had much smaller sample size, was less representative, service use was reported by parents only (not young people), and the reporting term was current (rather than 12 months) which would produce more unstable prevalence estimates. We selected the E-Risk Study over the MHCYP Study because the MHCYP Study had a much lower response rate which is likely to have led to non-response bias, and the reporting term was current (rather than 12 months) which would produce more unstable prevalence estimates. We selected the E-Risk Study over the REACH Study because the REACH Study had a smaller sample size, much lower response rate which is likely to have led to non-response bias, and the reporting term was current (rather than 12 months) which would produce more unstable prevalence estimates.

### F. Data source for mental health disorders and health service use: E-Risk Study, additional information

The Environmental Risk (E-Risk) Longitudinal Twin Study tracks the development of a birth cohort of 2,232 British children. The sample was drawn from a larger birth register of twins born in England and Wales in 1994-95.[40] Full details about the sample are reported elsewhere.[32] Briefly, the E-Risk sample was constructed in 1999-2000, when 1,116 families (93% of those eligible) with same-sex 5-year-old twins participated in home-visit assessments. This sample comprised 56% monozygotic (MZ) and 44% dizygotic (DZ) twin pairs; sex was evenly distributed within zygosity (49% male); and 90% of participants had a White ethnic background. Families were recruited to represent the UK population of families with newborns in the 1990s, on the basis of residential location throughout England and Wales and mother's age. Teenaged mothers with twins were over-selected to replace high-risk families who were selectively lost to the register through non-response. Older mothers having twins via assisted reproduction were under-selected to avoid an excess of well-educated older mothers. The study sample represents the full range of socioeconomic conditions in the UK, as reflected in the families' distribution on a neighbourhood-level socioeconomic index (ACORN [A Classification of Residential Neighbourhoods], developed by CACI Inc. for commercial use in Great Britain)[41]: 25.6% of E-Risk families live in "wealthy achiever" neighbourhoods compared to 25.3% nationwide; 5.3% vs. 11.6% live in "urban prosperity" neighbourhoods; 29.6% vs. 26.9% live in "comfortably off" neighbourhoods; 13.4% vs. 13.9% live in "moderate means" neighbourhoods; and 26.1% vs. 20.7% live in "hard-pressed" neighbourhoods. E-Risk underrepresents "urban prosperity" neighbourhoods because such households are likely to be childless.

Follow-up home visits were conducted when the children were aged 7 (98% participation), 10 (96% participation), 12 (96% participation), and 18 (93% participation) years old. Home visits at ages 5, 7, 10, and 12 years included assessments with participants as well as their mother (or primary caretaker); the home visit at age 18 included interviews only with participants. Each twin participant was assessed by a different interviewer. There were 2,066 study members who participated in the E-Risk assessments at age 18, and the proportions of MZ twins (55%), male same-sex twins (47%), and participants with a White ethnic background (91%) were almost identical to those found in the original sample at age 5. The average age of the twins at the time of assessment was 18.4 years (standard deviation [SD]=0.36); all interviews were conducted after the 18th birthday. The study sample at age 18 was equally distributed across IMD national deciles (figure below in appendix p15). There were no differences between those who did and did not take part at age 18 in terms of socioeconomic status assessed when the cohort was initially defined ($\chi2=0.86$, $p=0.65$), age-5 internalising or externalising symptoms ($t=0.40$, $p=0.69$ and $t=0.41$, $p=0.68$, respectively), or age-5 IQ scores ($t=0.98$, $p=0.33$).

We used data collected during the age-18 home visits in 2012-2013. Mental health disorders were assessed during private structured interviews that ascertained Diagnostic and Statistical Manual of Mental Disorders (DSM) criteria for depressive disorder (DSM-IV), generalised anxiety disorder (DSM-IV), post-traumatic stress disorder (PTSD; DSM-5), and attention-deficit/hyperactivity disorder (ADHD; DSM-5) in the past year.[9,11–13] Conduct disorder was assessed using self-completed computer-based surveys to ascertain DSM-IV criteria in the past year.[8] Health service use was assessed during interview by asking participants if they had seen a GP; a psychologist, counsellor, or therapist; or a psychiatrist for mental health problems in the past year. The prevalence of mental health disorders and health service use in the E-Risk Study, and their correlates, are provided below in appendix p16-17.

The Joint South London and Maudsley and the Institute of Psychiatry Research Ethics Committee approved each phase of the study. Parents gave informed consent and twins gave assent between 5 and 12 years and then informed consent at age 18.

**E-Risk Study IMD decile distribution**

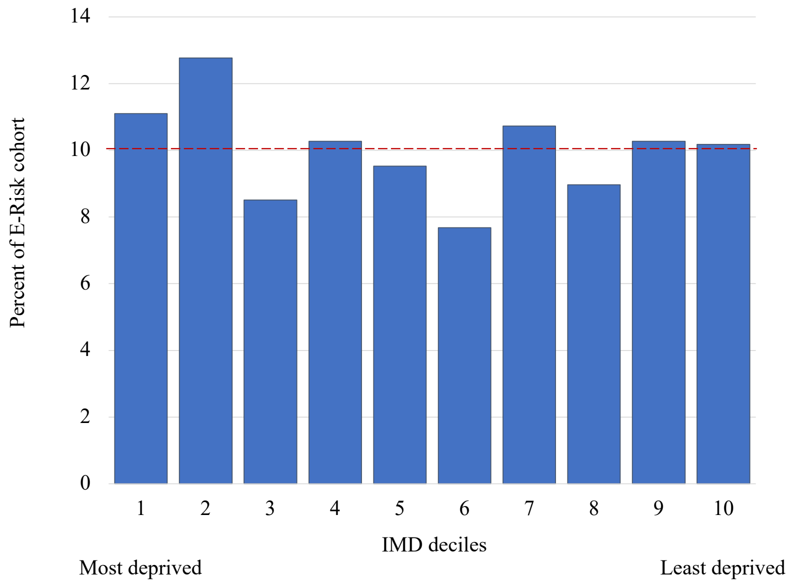

The figure shows that E-Risk families’ addresses at age 18 years are a close match to Index of Multiple Deprivation (IMD) national deciles; approximately 10% of the cohort fills each of IMD’s 10% bands for England.

### G. Data source for mental health disorders and health service use: E-Risk Study, mental health disorder prevalence

| | | | | Sex, Female vs Male | IMD quintile, increasing | Ethnicity, White vs Black, Mixed, Asian, and Other ethnicities |
|---|---|---|---|---|---|---|
| | n | N | % | OR (95% CI) | OR (95% CI) | OR (95% CI) |
| Any of the below disorders | 736 | 2,039 | 36.1 | 0.90 (0.74, 1.10) | **0.91 (0.85, 0.98)** | 0.94 (0.67, 1.33) |
| Depressive disorder | 414 | 2,063 | 20.1 | **1.57 (1.24, 2.00)** | 0.93 (0.85, 1.01) | 1.38 (0.89, 2.13) |
| Generalised anxiety disorder | 153 | 2,060 | 7.4 | **2.15 (1.47, 3.15)** | 0.97 (0.86, 1.09) | 1.55 (0.68, 3.53) |
| Post-traumatic stress disorder | 90 | 2,063 | 4.4 | **1.95 (1.21, 3.14)** | **0.84 (0.72, 0.98)** | 0.82 (0.40, 1.70) |
| Conduct disorder | 309 | 2,053 | 15.1 | **0.32 (0.24, 0.43)** | **0.90 (0.82, 1.00)** | 0.74 (0.46, 1.19) |
| Attention-deficit/hyperactivity disorder | 171 | 2,061 | 8.3 | 0.84 (0.60, 1.17) | 0.93 (0.82, 1.04) | 1.20 (0.66, 2.16) |
| Single disorder | 467 | 2,039 | 22.9 | **0.77 (0.62, 0.96)** | **0.92 (0.85, 1.00)** | 0.75 (0.54, 1.06) |
| Multiple disorders | 269 | 2,039 | 13.2 | 1.22 (0.92, 1.61) | 0.94 (0.85, 1.04) | 1.46 (0.87, 2.46) |

The table presents the number of participants who met criteria for each disorder (n), the number of participants who were assessed for the disorder (N), and the prevalence of the disorder (%; i.e., n/N as a percentage) during the year before age-18 assessment in the E-Risk Study. The table also presents odds ratios (OR) and their 95% confidence intervals (95% CI) for associations between socio-demographic characteristics and each disorder in E-Risk Study participants, calculated using generalised estimating equation logistic regression models, accounting for clustering within families. Index of Multiple Deprivation (IMD) national quintile is a measure of neighbourhood deprivation (1=most deprived; 5=least deprived), based on participants' home neighbourhood (lower-layer super output area) at age-18 follow up. Bold text indicates $p<0.05$.

### H. Data source for mental health disorders and health service use: E-Risk Study, health service use in participants who met criteria for a mental health disorder

| | | | | In participants with any disorder | | In participants with each disorder | | |
|---|---|---|---|---|---|---|---|---|
| | | | | Each disorder vs Other disorders | Multiple disorders vs Single disorder | Sex, Female vs Male | IMD quintile, increasing | Ethnicity, White vs Black, Mixed, Asian and other ethnicities |
| | n | N | % | OR (95%CI) | OR (95%CI) | OR (95%CI) | OR (95%CI) | OR (95%CI) |
| Any of the below disorders | 200 | 736 | 27.2 | - | - | **2.52 (1.78, 3.59)** | 1.09 (0.96, 1.24) | 1.47 (0.77, 2.78) |
| Depressive disorder | 158 | 413 | 38.3 | **3.97 (2.68, 5.89)** | - | **1.79 (1.16, 2.77)** | 1.14 (0.98, 1.33) | 1.08 (0.47, 2.48) |
| Generalised anxiety disorder | 80 | 153 | 52.3 | **4.10 (2.84, 5.91)** | - | 1.74 (0.89, 3.41) | 1.13 (0.87, 1.45) | 0.65 (0.17, 2.49) |
| Post-traumatic stress disorder | 37 | 90 | 41.1 | **1.90 (1.19, 3.04)** | - | 1.57 (0.61, 4.03) | 1.17 (0.83, 1.64) | 2.14 (0.41, 11.20) |
| Conduct disorder | 63 | 309 | 20.4 | **0.54 (0.38, 0.76)** | - | **1.96 (1.10, 3.48)** | 0.88 (0.70, 1.10) | 1.74 (0.67, 4.48) |
| Attention-deficit/hyperactivity disorder | 42 | 170 | 24.7 | 0.86 (0.58, 1.29) | - | 0.73 (0.36, 1.49) | **0.71 (0.55, 0.92)** | 2.04 (0.43, 9.68) |
| Single disorder | 89 | 467 | 19.1 | - | - | **3.39 (2.04, 5.65)** | 1.17 (0.99, 1.38) | 1.38 (0.62, 3.06) |
| Multiple disorders | 111 | 269 | 41.3 | - | **2.88 (2.07, 4.01)** | **1.75 (1.06, 2.90)** | 0.99 (0.82, 1.20) | 1.24 (0.43, 3.57) |

The table presents the number of participants who met criteria for each disorder (N) and reported using health services (n), and the prevalence of health service use in those who met criteria for each disorder (%; i.e., n/N as a percentage) during the year before age-18 assessment in the E-Risk Study. Additionally, the table presents odds ratios (OR) and their 95% confidence intervals (95% CI) for associations between each disorder vs other disorders and health service use, and associations between multiple disorders vs single disorder and health service use in E-Risk Study participants with any disorder. The table also presents odds ratios (OR) and their 95% confidence intervals (95% CI) for associations between socio-demographic characteristics and health service use in E-Risk Study participants with each disorder. Odds ratios and their confidence intervals were calculated using generalised estimating equation logistic regression models, accounting for clustering within families. Index of Multiple Deprivation (IMD) national quintile is a measure of neighbourhood deprivation (1=most deprived; 5=least deprived), based on participants' home neighbourhood (lower-layer super output area) at age-18 follow up. Bold text indicates $p<0.05$.

### I. Statistical analysis

When calculating the denominator for Samples 2 and 3 (the number of 17-year-olds who lived in South London in 2009-2024 and were estimated to have met criteria for a disorder during the past year, and the number who estimated to have also used any health services), we weighted according to sex and index of multiple deprivation (IMD) national quintile distributions in 17-year-old South Londoners (weight = number of South London 17-year-olds in sex-IMD quintile group / number of E-Risk participants in sex-IMD quintile group). We did not also weight by ethnicity in the main analyses because ethnicity data was less reliable owing to (1) the lack of annual population data on ethnicity and (2) the E-Risk sample containing fewer participants of Black, Asian, Mixed, and Other ethnicity than White ethnicity. Furthermore, sex and IMD national quintile were both associated with mental health disorders and health service use in the E-Risk Study, but ethnicity was not (appendix p16-17), and therefore it seemed appropriate to prioritise weighting by sex and IMD quintile. However, we did undertake sensitivity analyses also weighting by ethnicity. Because population ethnicity data was not available for 17-year-olds by sex and LSOA, when calculating weights by ethnicity, sex, and IMD quintile, we needed to consider ethnicity by broader age bands, and selected the narrowest available by sex and LSOA. For 2009-2015, we used ethnicity ratios for 0-24-year-olds from South London in the 2011 census.[42] For 2016-2024, we used ethnicity ratios for 16-17-year-olds from South London in the 2021 census.[43]

Analyses were conducted using Stata[44] and R.[45] Data were missing for a small proportion of diagnosed young people regarding their gender (n=18; 0.008% of the general population), neighbourhood (n=123, 0.06% of the general population), or ethnicity (n=417, 0.2% of the general population). Other gender was reported by a small number of diagnosed young people (n=53, 0.02%), which would not be sufficient for meaningful analysis and may be identifying, and is not included in population data, so we considered these data as missing. These young people were classed as undiagnosed for analyses that included their missing characteristics. In E-Risk Study participants who took part in the age-18 follow-up, data were missing for a small proportion regarding any disorder in the past year (n=27/2,066, 1.3%) and IMD quintile (n=197/2,066, 9.5%). Pairwise deletion was used for E-Risk data analyses that included these variables.

The analysis plan was pre-registered with the E-Risk Study team, and is available online: https://sites.duke.edu/moffittcaspiprojects/files/2026/04/Lewis_ERisk_mental-health-diagnoses-in-youth_final_29MAY2025.pdf. It was also pre-approved by the South London and Maudsley NHS Foundation Trust Clinical Record Interactive Search (CRIS) Oversight Committee.

### J. Diagnosis provision in the general population, Sample 1

| | N=223,404 | |
|---|---|---|
| | n | % |
| Any of the below disorders | 8,958 | 4.01 |
| Depressive disorder | 2,105 | 0.94 |
| Generalised anxiety disorder | 340 | 0.15 |
| Other anxiety disorder | 1,600 | 0.72 |
| Obsessive-compulsive disorder | 428 | 0.19 |
| Post-traumatic stress disorder | 703 | 0.31 |
| Other stress-related disorder | 669 | 0.30 |
| Dissociative or somatoform disorder | 142 | 0.06 |
| Eating disorder | 643 | 0.29 |
| Conduct disorder | 883 | 0.40 |
| Substance use disorder | 254 | 0.11 |
| Intellectual disability | 535 | 0.24 |
| Autistic spectrum disorder | 1,686 | 0.75 |
| Attention-deficit/hyperactivity disorder | 1,875 | 0.84 |
| Psychotic disorder | 315 | 0.14 |
| Mania or bipolar disorder | 98 | 0.04 |
| Single disorder | 6,187 | 2.77 |
| Multiple disorders | 2,771 | 1.24 |

The table presents the number (n) and rate (%) of diagnosis provision during the past year, in 17-year-olds from South London in 2009-2024.

### K. Diagnosis provision in the general population, Sample 1, by year

| | | 2009 | 2010 | 2011 | 2012 | 2013 | 2014 | 2015 | 2016 | 2017 | 2018 | 2019 | 2020 | 2021 | 2022 | 2023 | 2024 | Year, increasing |
|---|---|---|---|---|---|---|---|---|---|---|---|---|---|---|---|---|---|---|
| | N | 13,777 | 14,032 | 13,662 | 13,941 | 13,940 | 13,907 | 13,658 | 13,693 | 13,804 | 13,536 | 13,175 | 13,538 | 14,126 | 14,803 | 14,621 | 15,191 | OR (95% CI) |
| Any of the below disorders | n | 355 | 371 | 513 | 497 | 529 | 576 | 493 | 492 | 599 | 604 | 587 | 610 | 655 | 699 | 704 | 674 | **1.03 (1.03, 1.04)** |
| | % | 2.58 | 2.64 | 3.75 | 3.57 | 3.79 | 4.14 | 3.61 | 3.59 | 4.34 | 4.46 | 4.46 | 4.51 | 4.64 | 4.72 | 4.81 | 4.44 | |
| Depressive disorder | n | 99 | 99 | 119 | 137 | 149 | 170 | 120 | 132 | 158 | 143 | 139 | 131 | 139 | 137 | 124 | 109 | 1.00 (0.99, 1.01) |
| | % | 0.72 | 0.71 | 0.87 | 0.98 | 1.07 | 1.22 | 0.88 | 0.96 | 1.14 | 1.06 | 1.06 | 0.97 | 0.98 | 0.93 | 0.85 | 0.72 | |
| Generalised anxiety disorder | n | 5 | 10 | 22 | 12 | 18 | 20 | 20 | 16 | 21 | 32 | 24 | 20 | 20 | 37 | 34 | 29 | **1.07 (1.04, 1.09)** |
| | % | 0.04 | 0.07 | 0.16 | 0.09 | 0.13 | 0.14 | 0.15 | 0.12 | 0.15 | 0.24 | 0.18 | 0.15 | 0.14 | 0.25 | 0.23 | 0.19 | |
| Other anxiety disorder | n | 27 | 47 | 52 | 59 | 56 | 95 | 91 | 97 | 93 | 103 | 127 | 127 | 158 | 159 | 155 | 154 | **1.09 (1.08, 1.10)** |
| | % | 0.20 | 0.33 | 0.38 | 0.42 | 0.40 | 0.68 | 0.67 | 0.71 | 0.67 | 0.76 | 0.96 | 0.94 | 1.12 | 1.07 | 1.06 | 1.01 | |
| Obsessive-compulsive disorder | n | 11 | 17 | 20 | 23 | 27 | 30 | 22 | 13 | 30 | 34 | 27 | 23 | 29 | 45 | 43 | 34 | **1.05 (1.03, 1.08)** |
| | % | 0.08 | 0.12 | 0.15 | 0.16 | 0.19 | 0.22 | 0.16 | 0.09 | 0.22 | 0.25 | 0.20 | 0.17 | 0.21 | 0.30 | 0.29 | 0.22 | |
| Post-traumatic stress disorder | n | 12 | 26 | 31 | 34 | 33 | 32 | 25 | 36 | 46 | 47 | 49 | 55 | 52 | 59 | 89 | 77 | **1.09 (1.07, 1.11)** |
| | % | 0.09 | 0.19 | 0.23 | 0.24 | 0.24 | 0.23 | 0.18 | 0.26 | 0.33 | 0.35 | 0.37 | 0.41 | 0.37 | 0.40 | 0.61 | 0.51 | |
| Other stress-related disorder | n | 50 | 56 | 70 | 48 | 53 | 41 | 52 | 32 | 39 | 50 | 25 | 29 | 36 | 31 | 29 | 28 | **0.95 (0.93, 0.96)** |
| | % | 0.36 | 0.40 | 0.51 | 0.34 | 0.38 | 0.29 | 0.38 | 0.23 | 0.28 | 0.37 | 0.19 | 0.21 | 0.25 | 0.21 | 0.20 | 0.18 | |
| Dissociative or somatoform disorder | n | 3 | 2 | 8 | 12 | 5 | 13 | 6 | 8 | 11 | 9 | 13 | 12 | 10 | 11 | 12 | 7 | **1.04 (1.00, 1.08)** |
| | % | 0.02 | 0.01 | 0.06 | 0.09 | 0.04 | 0.09 | 0.04 | 0.06 | 0.08 | 0.07 | 0.10 | 0.09 | 0.07 | 0.07 | 0.08 | 0.05 | |
| Eating disorder | n | 27 | 22 | 28 | 24 | 27 | 35 | 23 | 33 | 44 | 50 | 50 | 44 | 53 | 77 | 53 | 53 | **1.07 (1.05, 1.09)** |
| | % | 0.20 | 0.16 | 0.20 | 0.17 | 0.19 | 0.25 | 0.17 | 0.24 | 0.32 | 0.37 | 0.38 | 0.33 | 0.38 | 0.52 | 0.36 | 0.35 | |
| Conduct disorder | n | 42 | 53 | 82 | 78 | 76 | 63 | 52 | 61 | 57 | 47 | 52 | 51 | 44 | 49 | 33 | 43 | **0.97 (0.95, 0.98)** |
| | % | 0.30 | 0.38 | 0.60 | 0.56 | 0.55 | 0.45 | 0.38 | 0.45 | 0.41 | 0.35 | 0.39 | 0.38 | 0.31 | 0.33 | 0.23 | 0.28 | |
| Substance use disorder | n | 31 | 21 | 29 | 23 | 15 | 14 | 11 | 16 | 17 | 15 | 16 | 11 | 13 | 2 | 13 | 7 | **0.92 (0.89, 0.95)** |
| | % | 0.23 | 0.15 | 0.21 | 0.16 | 0.11 | 0.10 | 0.08 | 0.12 | 0.12 | 0.11 | 0.12 | 0.08 | 0.09 | 0.01 | 0.09 | 0.05 | |
| Intellectual disability | n | 30 | 29 | 31 | 43 | 33 | 39 | 39 | 36 | 44 | 39 | 32 | 38 | 34 | 29 | 20 | 19 | **0.98 (0.96, 1.00)** |
| | % | 0.22 | 0.21 | 0.23 | 0.31 | 0.24 | 0.28 | 0.29 | 0.26 | 0.32 | 0.29 | 0.24 | 0.28 | 0.24 | 0.20 | 0.14 | 0.13 | |
| Autistic spectrum disorder | n | 36 | 36 | 59 | 64 | 92 | 96 | 83 | 85 | 110 | 136 | 130 | 152 | 143 | 147 | 151 | 166 | **1.08 (1.07, 1.09)** |
| | % | 0.26 | 0.26 | 0.43 | 0.46 | 0.66 | 0.69 | 0.61 | 0.62 | 0.80 | 1.00 | 0.99 | 1.12 | 1.01 | 0.99 | 1.03 | 1.09 | |
| Attention-deficit/hyperactivity disorder | n | 48 | 41 | 80 | 80 | 73 | 124 | 123 | 92 | 123 | 133 | 113 | 152 | 153 | 171 | 174 | 195 | **1.08 (1.07, 1.09)** |
| | % | 0.35 | 0.29 | 0.59 | 0.57 | 0.52 | 0.89 | 0.90 | 0.67 | 0.89 | 0.98 | 0.86 | 1.12 | 1.08 | 1.16 | 1.19 | 1.28 | |
| Psychotic disorder | n | 26 | 21 | 22 | 15 | 23 | 17 | 23 | 18 | 21 | 16 | 22 | 15 | 24 | 19 | 14 | 19 | 0.98 (0.96, 1.01) |
| | % | 0.19 | 0.15 | 0.16 | 0.11 | 0.16 | 0.12 | 0.17 | 0.13 | 0.15 | 0.12 | 0.17 | 0.11 | 0.17 | 0.13 | 0.10 | 0.13 | |
| Mania or bipolar disorder | n | 11 | 6 | 4 | 7 | 7 | 4 | 4 | 8 | 11 | 5 | 6 | 5 | 8 | 3 | 3 | 6 | 0.97 (0.93, 1.01) |
| | % | 0.08 | 0.04 | 0.03 | 0.05 | 0.05 | 0.03 | 0.03 | 0.06 | 0.08 | 0.04 | 0.05 | 0.04 | 0.06 | 0.02 | 0.02 | 0.04 | |
| Single disorder | n | 270 | 274 | 390 | 364 | 389 | 395 | 326 | 335 | 407 | 386 | 390 | 397 | 435 | 471 | 501 | 457 | **1.03 (1.02, 1.03)** |
| | % | 1.96 | 1.95 | 2.85 | 2.61 | 2.79 | 2.84 | 2.39 | 2.45 | 2.95 | 2.85 | 2.96 | 2.93 | 3.08 | 3.18 | 3.43 | 3.01 | |
| Multiple disorders | n | 85 | 97 | 123 | 133 | 140 | 181 | 167 | 157 | 192 | 218 | 197 | 213 | 220 | 228 | 203 | 217 | **1.05 (1.04, 1.06)** |
| | % | 0.62 | 0.69 | 0.90 | 0.95 | 1.00 | 1.30 | 1.22 | 1.15 | 1.39 | 1.61 | 1.50 | 1.57 | 1.56 | 1.54 | 1.39 | 1.43 | |

The table presents the number (n) and rate (%) of diagnosis provision during the past year, in 17-year-olds from South London in 2009-2024, by year. The table also presents odds ratios (OR) and their 95% confidence intervals (95% CI) for associations between year and diagnosis provision in these young people. Bold text indicates p<0.05.

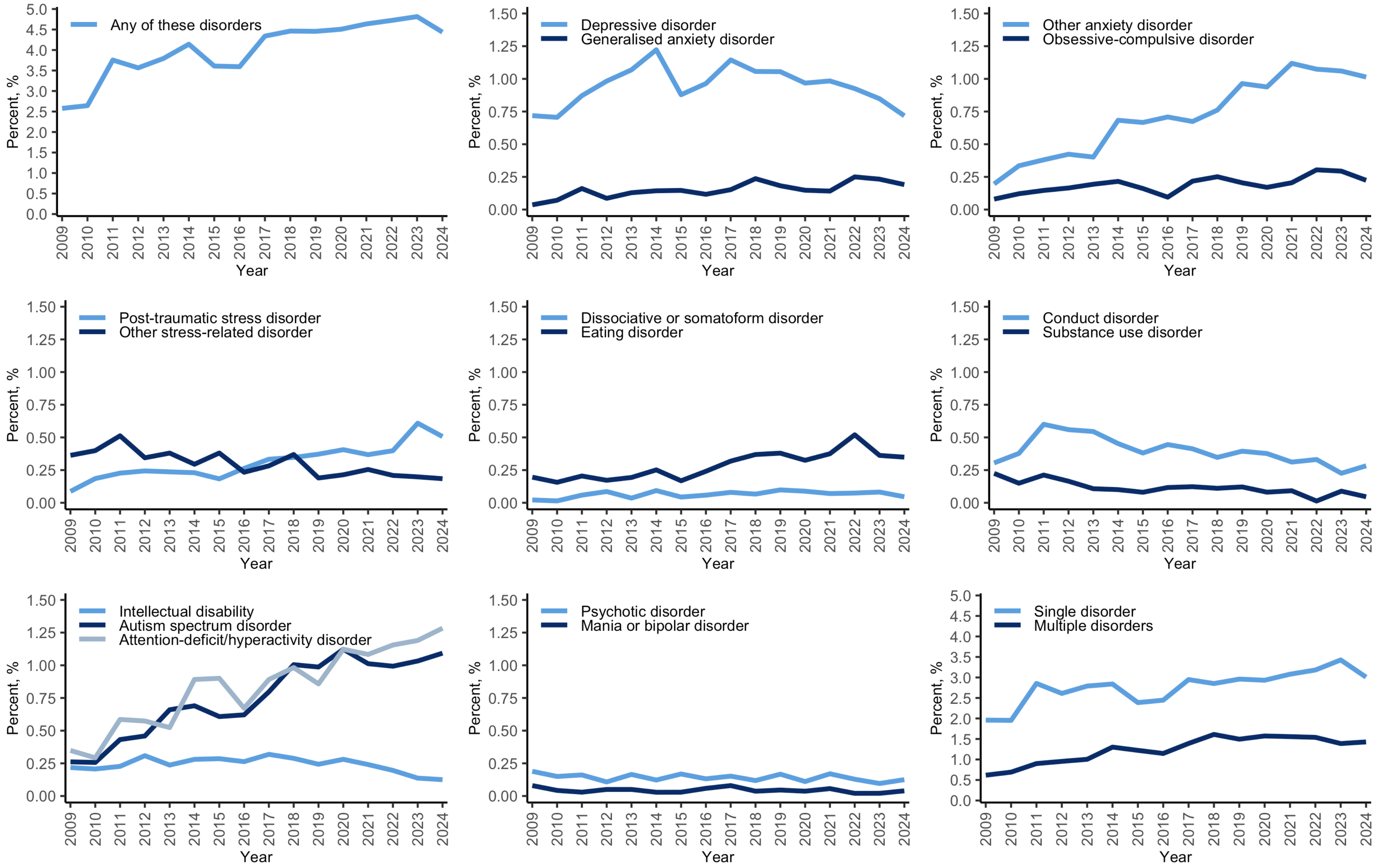

The plot presents the rate of diagnosis provision during the past year, in 17-year-olds from South London in 2009-2024, by year.

### L. Diagnosis provision in the general population, Sample 1, by gender

| | | Male N=113,877 | Female N=109,527 | Female vs Male OR (95% CI) |
|---|---|---|---|---|
| Any of the below disorders | n | 4,113 | 4,774 | **1.22 (1.17, 1.27)** |
| | % | 3.61 | 4.36 | |
| Depressive disorder | n | 600 | 1,490 | **2.60 (2.37, 2.86)** |
| | % | 0.53 | 1.36 | |
| Generalised anxiety disorder | n | 97 | 237 | **2.54 (2.01, 3.22)** |
| | % | 0.09 | 0.22 | |
| Other anxiety disorder | n | 478 | 1,102 | **2.41 (2.17, 2.69)** |
| | % | 0.42 | 1.01 | |
| Obsessive-compulsive disorder | n | 179 | 247 | **1.44 (1.18, 1.74)** |
| | % | 0.16 | 0.23 | |
| Post-traumatic stress disorder | n | 245 | 451 | **1.92 (1.64, 2.24)** |
| | % | 0.22 | 0.41 | |
| Other stress-related disorder | n | 210 | 453 | **2.25 (1.91, 2.65)** |
| | % | 0.18 | 0.41 | |
| Dissociative or somatoform disorder | n | 39 | 100 | **2.67 (1.84, 3.86)** |
| | % | 0.03 | 0.09 | |
| Eating disorder | n | 47 | 590 | **13.12 (9.74, 17.66)** |
| | % | 0.04 | 0.54 | |
| Conduct disorder | n | 617 | 264 | **0.44 (0.38, 0.51)** |
| | % | 0.54 | 0.24 | |
| Substance use disorder | n | 170 | 84 | **0.51 (0.40, 0.67)** |
| | % | 0.15 | 0.08 | |
| Intellectual disability | n | 390 | 145 | **0.39 (0.32, 0.47)** |
| | % | 0.34 | 0.13 | |
| Autistic spectrum disorder | n | 1,171 | 494 | **0.44 (0.39, 0.48)** |
| | % | 1.03 | 0.45 | |
| Attention-deficit/hyperactivity disorder | n | 1,476 | 394 | **0.27 (0.25, 0.31)** |
| | % | 1.30 | 0.36 | |
| Psychotic disorder | n | 190 | 125 | **0.68 (0.55, 0.86)** |
| | % | 0.17 | 0.11 | |
| Mania or bipolar disorder | n | 37 | 61 | **1.71 (1.14, 2.58)** |
| | % | 0.03 | 0.06 | |
| Single disorder | n | 2,613 | 3,523 | **1.42 (1.34, 1.49)** |
| | % | 2.29 | 3.22 | |
| Multiple disorders | n | 1,500 | 1,251 | **0.87 (0.80, 0.93)** |
| | % | 1.32 | 1.14 | |

The table presents the number (n) and rate (%) of diagnosis provision during the past year, in 17-year-olds from London in 2009-2024, by gender. The table also presents odds ratios (OR) and their 95% confidence intervals (95% CI) for associations between gender and diagnosis provision in these young people. Bold text indicates p<0.05.

### M. Diagnosis provision in the general population, Sample 1, by neighbourhood deprivation (IMD quintile)

| | | 1 (most deprived) N=65,554 | 2 N=85,933 | 3 N=42,310 | 4 N=19,164 | 5 (least deprived) N=10,443 | IMD quintile, increasing OR (95% CI) |
|---|---|---|---|---|---|---|---|
| Any of the below disorders | n | 2,611 | 3,347 | 1,830 | 734 | 313 | **0.98 (0.96, 1.00)** |
| | % | 3.98 | 3.89 | 4.33 | 3.83 | 3.00 | |
| Depressive disorder | n | 568 | 802 | 455 | 188 | 68 | 1.01 (0.98, 1.05) |
| | % | 0.87 | 0.93 | 1.08 | 0.98 | 0.65 | |
| Generalised anxiety disorder | n | 72 | 121 | 88 | 36 | 22 | **1.21 (1.10, 1.32)** |
| | % | 0.11 | 0.14 | 0.21 | 0.19 | 0.21 | |
| Other anxiety disorder | n | 406 | 599 | 354 | 159 | 66 | **1.07 (1.03, 1.12)** |
| | % | 0.62 | 0.70 | 0.84 | 0.83 | 0.63 | |
| Obsessive-compulsive disorder | n | 87 | 147 | 109 | 60 | 21 | **1.23 (1.14, 1.34)** |
| | % | 0.13 | 0.17 | 0.26 | 0.31 | 0.20 | |
| Post-traumatic stress disorder | n | 189 | 292 | 143 | 50 | 19 | 0.96 (0.90, 1.03) |
| | % | 0.29 | 0.34 | 0.34 | 0.26 | 0.18 | |
| Other stress-related disorder | n | 209 | 246 | 138 | 47 | 13 | **0.91 (0.84, 0.98)** |
| | % | 0.32 | 0.29 | 0.33 | 0.25 | 0.12 | |
| Dissociative or somatoform disorder | n | 30 | 56 | 35 | 12 | 7 | 1.13 (0.97, 1.30) |
| | % | 0.05 | 0.07 | 0.08 | 0.06 | 0.07 | |
| Eating disorder | n | 110 | 206 | 168 | 110 | 47 | **1.39 (1.30, 1.47)** |
| | % | 0.17 | 0.24 | 0.40 | 0.57 | 0.45 | |
| Conduct disorder | n | 323 | 326 | 151 | 38 | 19 | **0.79 (0.74, 0.85)** |
| | % | 0.49 | 0.38 | 0.36 | 0.20 | 0.18 | |
| Substance use disorder | n | 82 | 99 | 47 | 12 | 3 | **0.82 (0.72, 0.93)** |
| | % | 0.13 | 0.12 | 0.11 | 0.06 | 0.03 | |
| Intellectual disability | n | 231 | 177 | 76 | 29 | 14 | **0.74 (0.68, 0.81)** |
| | % | 0.35 | 0.21 | 0.18 | 0.15 | 0.13 | |
| Autistic spectrum disorder | n | 558 | 599 | 329 | 116 | 69 | **0.93 (0.89, 0.97)** |
| | % | 0.85 | 0.70 | 0.78 | 0.61 | 0.66 | |
| Attention-deficit/hyperactivity disorder | n | 640 | 694 | 341 | 112 | 63 | **0.88 (0.84, 0.92)** |
| | % | 0.98 | 0.81 | 0.81 | 0.58 | 0.60 | |
| Psychotic disorder | n | 87 | 144 | 57 | 20 | 3 | **0.88 (0.79, 0.98)** |
| | % | 0.13 | 0.17 | 0.13 | 0.10 | 0.03 | |
| Mania or bipolar disorder | n | 35 | 34 | 19 | 9 | 0 | 0.85 (0.69, 1.03) |
| | % | 0.05 | 0.04 | 0.04 | 0.05 | 0.00 | |
| Single disorder | n | 1,727 | 2,365 | 1,268 | 514 | 218 | 0.99 (0.97, 1.02) |
| | % | 2.63 | 2.75 | 3.00 | 2.68 | 2.09 | |
| Multiple disorders | n | 884 | 982 | 562 | 220 | 95 | **0.95 (0.92, 0.98)** |
| | % | 1.35 | 1.14 | 1.33 | 1.15 | 0.91 | |

The table presents the number (n) and rate (%) of diagnosis provision during the past year, in 17-year-olds from South London in 2009-2024, by Index of Multiple Deprivation (IMD) national quintile. The table also presents odds ratios (OR) and their 95% confidence intervals (95% CI) for associations between IMD national quintile and diagnosis provision in these young people. Bold text indicates $p<0.05$.

### N. Diagnosis provision in the general population, Sample 1, by ethnicity

| | | White N=75,995 | Black N=82,151 | Mixed N=27,186 | Asian N=26,589 | Other ethnicities N=11,483 | Black, Mixed, Asian, and Other ethnicities N=147,409 | White vs Black, Mixed, Asian, or Other ethnicity OR (0.95 CI) |
|---|---|---|---|---|---|---|---|---|
| Any of the below disorders | n | 4,375 | 2,232 | 992 | 531 | 411 | 4,166 | **2.10 (2.01, 2.19)** |
| | % | 5.76 | 2.72 | 3.65 | 2.00 | 3.58 | 2.83 | |
| Depressive disorder | n | 1,010 | 469 | 247 | 141 | 126 | 983 | **2.01 (1.84, 2.19)** |
| | % | 1.33 | 0.57 | 0.91 | 0.53 | 1.10 | 0.67 | |
| Generalised anxiety disorder | n | 214 | 38 | 37 | 18 | 13 | 106 | **3.92 (3.11, 4.95)** |
| | % | 0.28 | 0.05 | 0.14 | 0.07 | 0.11 | 0.07 | |
| Other anxiety disorder | n | 896 | 269 | 175 | 99 | 58 | 601 | **2.91 (2.63, 3.23)** |
| | % | 1.18 | 0.33 | 0.64 | 0.37 | 0.51 | 0.41 | |
| Obsessive-compulsive disorder | n | 253 | 59 | 46 | 37 | 15 | 157 | **3.13 (2.57, 3.82)** |
| | % | 0.33 | 0.07 | 0.17 | 0.14 | 0.13 | 0.11 | |
| Post-traumatic stress disorder | n | 240 | 203 | 74 | 89 | 65 | 431 | 1.08 (0.92, 1.27) |
| | % | 0.32 | 0.25 | 0.27 | 0.33 | 0.57 | 0.29 | |
| Other stress-related disorder | n | 272 | 208 | 71 | 49 | 28 | 356 | **1.48 (1.27, 1.74)** |
| | % | 0.36 | 0.25 | 0.26 | 0.18 | 0.24 | 0.24 | |
| Dissociative or somatoform disorder | n | 67 | 27 | 16 | 13 | 13 | 69 | **1.88 (1.35, 2.64)** |
| | % | 0.09 | 0.03 | 0.06 | 0.05 | 0.11 | 0.05 | |
| Eating disorder | n | 430 | 67 | 64 | 33 | 15 | 179 | **4.68 (3.93, 5.57)** |
| | % | 0.57 | 0.08 | 0.24 | 0.12 | 0.13 | 0.12 | |
| Conduct disorder | n | 439 | 259 | 112 | 23 | 31 | 425 | **2.01 (1.76, 2.30)** |
| | % | 0.58 | 0.32 | 0.41 | 0.09 | 0.27 | 0.29 | |
| Substance use disorder | n | 97 | 87 | 37 | 9 | 14 | 147 | 1.28 (0.99, 1.65) |
| | % | 0.13 | 0.11 | 0.14 | 0.03 | 0.12 | 0.10 | |
| Intellectual disability | n | 204 | 219 | 43 | 36 | 21 | 319 | **1.24 (1.04, 1.48)** |
| | % | 0.27 | 0.27 | 0.16 | 0.14 | 0.18 | 0.22 | |
| Autistic spectrum disorder | n | 814 | 498 | 180 | 92 | 61 | 831 | **1.91 (1.73, 2.10)** |
| | % | 1.07 | 0.61 | 0.66 | 0.35 | 0.53 | 0.56 | |
| Attention-deficit/hyperactivity disorder | n | 1,057 | 442 | 234 | 46 | 63 | 785 | **2.63 (2.40, 2.89)** |
| | % | 1.39 | 0.54 | 0.86 | 0.17 | 0.55 | 0.53 | |
| Psychotic disorder | n | 61 | 178 | 24 | 31 | 17 | 250 | **0.47 (0.36, 0.63)** |
| | % | 0.08 | 0.22 | 0.09 | 0.12 | 0.15 | 0.17 | |
| Mania or bipolar disorder | n | 27 | 36 | 21 | 11 | 3 | 71 | 0.74 (0.47, 1.15) |
| | % | 0.04 | 0.04 | 0.08 | 0.04 | 0.03 | 0.05 | |
| Single disorder | n | 2,940 | 1,554 | 669 | 365 | 303 | 2,891 | **2.01 (1.91, 2.12)** |
| | % | 3.87 | 1.89 | 2.46 | 1.37 | 2.64 | 1.96 | |
| Multiple disorders | n | 1,435 | 678 | 323 | 166 | 108 | 1,275 | **2.21 (2.04, 2.38)** |
| | % | 1.89 | 0.83 | 1.19 | 0.62 | 0.94 | 0.86 | |

The table presents the number (n) and rate (%) of diagnosis provision during the past year, in 17-year-olds from South London in 2009-2024, by ethnicity. The table also presents odds ratios (OR) and their 95% confidence intervals (95% CI) for associations between ethnicity and diagnosis provision in these young people. Bold text indicates p<0.05.

#### O. Diagnosis provision in those estimated to have met criteria for a disorder, Sample 2

| | n | N | % | Each disorder vs Other disorders OR (95% CI) | Multiple disorders vs Single disorder OR (95% CI) |
|---|---|---|---|---|---|
| Any of the below disorders | 5,132 | 81,050 | 6.3 | - | - |
| Depressive disorder | 2,105 | 45,755 | 4.6 | **0.51 (0.49, 0.54)** | - |
| Generalised anxiety disorder | 340 | 17,131 | 2.0 | **0.25 (0.22, 0.28)** | - |
| Post-traumatic stress disorder | 703 | 10,747 | 6.5 | 1.04 (0.96, 1.13) | - |
| Conduct disorder | 883 | 33,560 | 2.6 | **0.27 (0.26, 0.30)** | - |
| Attention-deficit/hyperactivity disorder | 1,875 | 19,102 | 9.8 | **1.96 (1.85, 2.08)** | - |
| Single disorder | 4,384 | 51,433 | 8.5 | - | - |
| Multiple disorders | 748 | 29,617 | 2.5 | - | **0.28 (0.26, 0.30)** |

The table presents the number of 17-year-olds from South London in 2009-2024 who were diagnosed with each disorder (n), the number who were estimated to have met criteria for the disorder (N), and the rate of diagnosis provision in those estimated to have met criteria for the disorder (%; i.e., n/N as a percentage) during the past year. The table also presents odds ratios (OR) and their 95% confidence intervals (95% CI) for associations between each disorder vs other disorders and diagnosis provision, and associations between multiple disorders vs single disorder and diagnosis provision in these young people estimated to have any disorder. Bold text indicates $p<0.05$. These confidence intervals (and p value indications) do not account for possible error of N in relation to the true number who met criteria for the disorder.

### P. Diagnosis provision in those estimated to have met criteria for a disorder, Sample 2, sensitivity analysis also weighted by ethnicity

| | n | N | % | **Each disorder vs Other disorders** OR (95% CI) | **Multiple disorders vs Single disorder** OR (95% CI) |
|---|---|---|---|---|---|
| Any of the below disorders | 5,132 | 86,844 | 5.9 | - | - |
| Depressive disorder | 2,105 | 43,336 | 4.9 | **0.68 (0.64, 0.72)** | - |
| Generalised anxiety disorder | 340 | 14,047 | 2.4 | **0.35 (0.31, 0.39)** | - |
| Post-traumatic stress disorder | 703 | 14,232 | 4.9 | **0.80 (0.74, 0.87)** | - |
| Conduct disorder | 883 | 41,716 | 2.1 | **0.21 (0.19, 0.22)** | - |
| Attention-deficit/hyperactivity disorder | 1,875 | 19,780 | 9.5 | **2.05 (1.93, 2.18)** | - |
| Single disorder | 4,384 | 57,018 | 7.7 | - | - |
| Multiple disorders | 748 | 29,825 | 2.5 | - | **0.31 (0.29, 0.33)** |

The table presents the number of 17-year-olds from South London in 2009-2024 who were diagnosed with each disorder (n), the number who were estimated to have met criteria for the disorder (N), and the rate of diagnosis provision in those estimated to have met criteria for the disorder (%; i.e., n/N as a percentage) during the past year. N was weighted by ethnicity as well as gender and neighbourhood deprivation. The table also presents odds ratios (OR) and their 95% confidence intervals (95% CI) for associations between each disorder vs other disorders and diagnosis provision, and associations between multiple disorders vs single disorder and diagnosis provision in these young people estimated to have any disorder. Bold text indicates $p<0.05$. These confidence intervals (and p value indications) do not account for possible error of N in relation to the true number who met criteria for the disorder.

### Q. Diagnosis provision in those estimated to have met criteria for a disorder, Sample 2, by gender

| | | Male | Female | Female vs Male OR (95% CI) |
|---|---|---|---|---|
| Any of the below disorders | n | 2,553 | 2,547 | 1.05 (0.99, 1.11) |
| | N | 41,544 | 39,506 | |
| | % | 6.1 | 6.4 | |
| Depressive disorder | n | 600 | 1,490 | **1.74 (1.58, 1.91)** |
| | N | 18,580 | 27,176 | |
| | % | 3.2 | 5.5 | |
| Generalised anxiety disorder | n | 97 | 237 | **1.34 (1.06, 1.70)** |
| | N | 6,056 | 11,075 | |
| | % | 1.6 | 2.1 | |
| Post-traumatic stress disorder | n | 245 | 451 | 0.92 (0.78, 1.08) |
| | N | 3,592 | 7,154 | |
| | % | 6.8 | 6.3 | |
| Conduct disorder | n | 617 | 264 | 1.15 (0.99, 1.33) |
| | N | 24,416 | 9,145 | |
| | % | 2.5 | 2.9 | |
| Attention-deficit/hyperactivity disorder | n | 1,476 | 394 | **0.31 (0.27, 0.34)** |
| | N | 10,688 | 8,414 | |
| | % | 13.8 | 4.7 | |
| Single disorder | n | 2,088 | 2,267 | **1.32 (1.24, 1.40)** |
| | N | 27,911 | 23,522 | |
| | % | 7.5 | 9.6 | |
| Multiple disorders | n | 465 | 280 | **0.50 (0.43, 0.59)** |
| | N | 13,633 | 15,984 | |
| | % | 3.4 | 1.8 | |

The table presents the number of 17-year-olds from South London in 2009-2024 who were diagnosed with each disorder (n), the number who were estimated to have met criteria for the disorder (N), and the rate of diagnosis provision in those estimated to have met criteria for the disorder (%; i.e., n/N as a percentage) during the past year, by gender. The table also presents odds ratios (OR) and their 95% confidence intervals (95% CI) for associations between gender and diagnosis provision in these young people estimated to have each disorder. Bold text indicates $p<0.05$. These confidence intervals (and p value indications) do not account for possible error of N in relation to the true number who met criteria for the disorder.

**R. Diagnosis provision in those estimated to have met criteria for a disorder, Sample 2, by neighbourhood deprivation (IMD quintile)**

| | | 1 (most deprived) | 2 | 3 | 4 | 5 (least deprived) | IMD quintile, increasing OR (95% CI) |
|---|---|---|---|---|---|---|---|
| Any of the below disorders | n | 1,539 | 1,929 | 1,037 | 387 | 166 | 1.01 (0.99, 1.04) |
| | N | 26,283 | 30,134 | 15,385 | 5,804 | 3,444 | |
| | % | 5.9 | 6.4 | 6.7 | 6.7 | 4.8 | |
| Depressive disorder | n | 568 | 802 | 455 | 188 | 68 | **1.08 (1.04, 1.12)** |
| | N | 14,166 | 17,951 | 8,727 | 2,932 | 1,979 | |
| | % | 4.0 | 4.5 | 5.2 | 6.4 | 3.4 | |
| Generalised anxiety disorder | n | 72 | 121 | 88 | 36 | 22 | **1.21 (1.09, 1.33)** |
| | N | 4,493 | 6,425 | 4,321 | 1,294 | 597 | |
| | % | 1.6 | 1.9 | 2.0 | 2.8 | 3.7 | |
| Post-traumatic stress disorder | n | 189 | 292 | 143 | 50 | 19 | **1.12 (1.04, 1.20)** |
| | N | 3,488 | 4,593 | 1,551 | 863 | 251 | |
| | % | 5.4 | 6.4 | 9.2 | 5.8 | 7.6 | |
| Conduct disorder | n | 323 | 326 | 151 | 38 | 19 | **0.85 (0.79, 0.91)** |
| | N | 11,174 | 11,825 | 7,050 | 2,110 | 1,402 | |
| | % | 2.9 | 2.8 | 2.1 | 1.8 | 1.4 | |
| Attention-deficit/hyperactivity disorder | n | 640 | 694 | 341 | 112 | 63 | **0.93 (0.89, 0.98)** |
| | N | 5,837 | 8,023 | 2,869 | 1,716 | 657 | |
| | % | 11.0 | 8.7 | 11.9 | 6.5 | 9.6 | |
| Single disorder | n | 1,286 | 1,631 | 909 | 352 | 143 | **1.06 (1.03, 1.09)** |
| | N | 17,797 | 18,457 | 9,347 | 3,498 | 2,334 | |
| | % | 7.2 | 8.8 | 9.7 | 10.1 | 6.1 | |
| Multiple disorders | n | 253 | 298 | 128 | 35 | 23 | **0.85 (0.79, 0.91)** |
| | N | 8,486 | 11,677 | 6,038 | 2,306 | 1,110 | |
| | % | 3.0 | 2.6 | 2.1 | 1.5 | 2.1 | |

The table presents the number of 17-year-olds from South London in 2009-2024 who were diagnosed with each disorder (n), the number who were estimated to have met criteria for the disorder (N), and the rate of diagnosis provision in those estimated to have met criteria for the disorder (%; i.e., n/N as a percentage) during the past year, by Index of Multiple Deprivation (IMD) national quintile. The table also presents odds ratios (OR) and their 95% confidence intervals (95% CI) for associations between IMD national quintile and diagnosis provision in these young people estimated to have each disorder. Bold text indicates $p<0.05$. These confidence intervals (and p value indications) do not account for possible error of N in relation to the true number who met criteria for the disorder.

### S. Diagnosis provision in those estimated to have met criteria for a disorder, Sample 2, by ethnicity

| | | White | Black, Mixed, Asian, and Other ethnicities | White vs Black, Mixed, Asian, or Other ethnicity OR (95% CI) |
|---|---|---|---|---|
| Any of the below disorders | n | 2,544 | 2,383 | |
| | N | 27,459 | 55,303 | **2.27 (2.14, 2.40)** |
| | % | 9.3 | 4.3 | |
| Depressive disorder | n | 1,010 | 983 | |
| | N | 15,884 | 24,960 | **1.66 (1.51, 1.81)** |
| | % | 6.4 | 3.9 | |
| Generalised anxiety disorder | n | 214 | 106 | |
| | N | 6,132 | 6,305 | **2.11 (1.67, 2.68)** |
| | % | 3.5 | 1.7 | |
| Post-traumatic stress disorder | n | 240 | 431 | |
| | N | 3,566 | 8,563 | **1.36 (1.16, 1.60)** |
| | % | 6.7 | 5.0 | |
| Conduct disorder | n | 439 | 425 | |
| | N | 11,168 | 26,213 | **2.48 (2.17, 2.84)** |
| | % | 3.9 | 1.6 | |
| Attention-deficit/hyperactivity disorder | n | 1,057 | 785 | |
| | N | 6,575 | 11,345 | **2.58 (2.34, 2.84)** |
| | % | 16.1 | 6.9 | |
| Single disorder | n | 2,143 | 2,047 | |
| | N | 17,086 | 40,628 | **2.70 (2.54, 2.88)** |
| | % | 12.5 | 5.0 | |
| Multiple disorders | n | 401 | 336 | |
| | N | 10,373 | 14,675 | **1.72 (1.48, 1.99)** |
| | % | 3.9 | 2.3 | |

The table presents the number of 17-year-olds from South London in 2009-2024 who were diagnosed with each disorder (n), the number who were estimated to have met criteria for the disorder (N), and the rate of diagnosis provision in those estimated to have met criteria for the disorder (%; i.e., n/N as a percentage) during the past year, by ethnicity. The table also presents odds ratios (OR) and their 95% confidence intervals (95% CI) for associations between ethnicity and diagnosis provision in these young people estimated to have each disorder. Bold text indicates $p<0.05$. These confidence intervals (and p value indications) do not account for possible error of N in relation to the true number who met criteria for the disorder.

**T. Diagnosis provision in those estimated to have met criteria for a disorder and used health services, Sample 3**

| | n | N | % | Each disorder vs Other disorders OR (95% CI) | Multiple disorders vs Single disorder OR (95% CI) |
|---|---|---|---|---|---|
| Any of the below disorders | 5,132 | 21,598 | 23.8 | - | - |
| Depressive disorder | 2,105 | 17,243 | 12.2 | **0.06 (0.06, 0.07)** | - |
| Generalised anxiety disorder | 340 | 8,778 | 3.9 | **0.07 (0.06, 0.08)** | - |
| Post-traumatic stress disorder | 703 | 3,983 | 17.6 | **0.64 (0.58, 0.70)** | - |
| Conduct disorder | 883 | 7,359 | 12.0 | **0.32 (0.30, 0.35)** | - |
| Attention-deficit/hyperactivity disorder | 1,875 | 5,454 | 34.4 | **2.07 (1.94, 2.22)** | - |
| Single disorder | 4,384 | 9,219 | 47.6 | - | - |
| Multiple disorders | 748 | 12,379 | 6.0 | - | **0.07 (0.07, 0.08)** |

The table presents the number of 17-year-olds from South London in 2009-2024 who were diagnosed with each disorder (n), the number who were estimated to have met criteria for the disorder and used health services (N), and the rate of diagnosis provision in those estimated to have met criteria for the disorder and used health services (%; i.e., n/N as a percentage) during the past year. The table also presents odds ratios (OR) and their 95% confidence intervals (95% CI) for associations between each disorder vs other disorders and diagnosis provision, and associations between multiple disorders vs single disorder and diagnosis provision in these young people estimated to have any disorder. Bold text indicates $p<0.05$. These confidence intervals (and p value indications) do not account for possible error of N in relation to the true number who met criteria for the disorder and used health services.

**U. Diagnosis provision in those estimated to have met criteria for a disorder and used health services, Sample 3, sensitivity analysis also weighted by ethnicity**

| | n | N | % | Each disorder vs Other disorders OR (95% CI) | Multiple disorders vs Single disorder OR (95% CI) |
|---|---|---|---|---|---|
| Any of the below disorders | 5,132 | 23,600 | 21.7 | - | - |
| Depressive disorder | 2,105 | 18,614 | 11.3 | **0.08 (0.08, 0.09)** | - |
| Generalised anxiety disorder | 340 | 9,460 | 3.6 | **0.07 (0.06, 0.08)** | - |
| Post-traumatic stress disorder | 703 | 6,299 | 11.2 | **0.37 (0.34, 0.40)** | - |
| Conduct disorder | 883 | 10,773 | 8.2 | **0.18 (0.17, 0.19)** | - |
| Attention-deficit/hyperactivity disorder | 1,875 | 4,324 | 43.4 | **3.77 (3.51, 4.04)** | - |
| Single disorder | 4,384 | 10,186 | 43.0 | - | - |
| Multiple disorders | 748 | 13,414 | 5.6 | - | **0.08 (0.07, 0.08)** |

The table presents the number of 17-year-olds from South London in 2009-2024 who were diagnosed with each disorder (n), the number who were estimated to have met criteria for the disorder and used health services (N), and the rate of diagnosis provision in those estimated to have met criteria for the disorder and used health services (%; i.e., n/N as a percentage) during the past year. N was weighted by ethnicity as well as gender and neighbourhood deprivation. The table also presents odds ratios (OR) and their 95% confidence intervals (95% CI) for associations between each disorder vs other disorders and diagnosis provision, and associations between multiple disorders vs single disorder and diagnosis provision in these young people estimated to have any disorder. Bold text indicates $p<0.05$. These confidence intervals (and p value indications) do not account for possible error of N in relation to the true number who met criteria for the disorder and used health services.

### V. Diagnosis provision in those estimated to have met criteria for a disorder and used health services, Sample 3, by gender

| | | Male | Female | Female vs Male OR (95% CI) |
|---|---|---|---|---|
| Any of the below disorders | n | 2,553 | 2,547 | **0.50 (0.47, 0.53)** |
| | N | 8,049 | 13,549 | |
| | % | 31.7 | 18.8 | |
| Depressive disorder | n | 600 | 1,490 | **1.14 (1.03, 1.26)** |
| | N | 5,374 | 11,869 | |
| | % | 11.2 | 12.6 | |
| Generalised anxiety disorder | n | 97 | 237 | 1.14 (0.89, 1.44) |
| | N | 2,776 | 6,003 | |
| | % | 3.5 | 3.9 | |
| Post-traumatic stress disorder | n | 245 | 451 | 0.92 (0.77, 1.09) |
| | N | 1,339 | 2,644 | |
| | % | 18.3 | 17.1 | |
| Conduct disorder | n | 617 | 264 | **0.80 (0.68, 0.93)** |
| | N | 4,831 | 2,528 | |
| | % | 12.8 | 10.4 | |
| Attention-deficit/hyperactivity disorder | n | 1,476 | 394 | **0.30 (0.26, 0.34)** |
| | N | 3,373 | 2,081 | |
| | % | 43.8 | 18.9 | |
| Single disorder | n | 2,088 | 2,267 | **0.28 (0.26, 0.31)** |
| | N | 3,089 | 6,130 | |
| | % | 67.6 | 37.0 | |
| Multiple disorders | n | 465 | 280 | **0.38 (0.33, 0.44)** |
| | N | 4,960 | 7,419 | |
| | % | 9.4 | 3.8 | |

The table presents the number of 17-year-olds from South London in 2009-2024 who were diagnosed with each disorder (n), the number who were estimated to have met criteria for the disorder and used health services (N), and the rate of diagnosis provision in those estimated to have met criteria for the disorder and used health services (%; i.e., n/N as a percentage) during the past year, by gender. The table also presents odds ratios (OR) and their 95% confidence intervals (95% CI) for associations between gender and diagnosis provision in these young people estimated to have each disorder. Bold text indicates $p<0.05$. These confidence intervals (and p value indications) do not account for possible error of N in relation to the true number who met criteria for the disorder and used health services.

**W. Diagnosis provision in those estimated to have met criteria for a disorder and used health services, Sample 3, by neighbourhood deprivation (IMD quintile)**

| | | 1 (most deprived) | 2 | 3 | 4 | 5 (least deprived) | IMD quintile, increasing OR (95% CI) |
|---|---|---|---|---|---|---|---|
| Any of the below disorders | n | 1,539 | 1,929 | 1,037 | 387 | 166 | **0.93 (0.90, 0.95)** |
| | N | 6,341 | 7,924 | 4,549 | 1,782 | 1,003 | |
| | % | 24.3 | 24.3 | 22.8 | 21.7 | 16.5 | |
| Depressive disorder | n | 568 | 802 | 455 | 188 | 68 | 0.96 (0.92, 1.00) |
| | N | 4,413 | 6,857 | 3,865 | 1,241 | 868 | |
| | % | 12.9 | 11.7 | 11.8 | 15.2 | 7.8 | |
| Generalised anxiety disorder | n | 72 | 121 | 88 | 36 | 22 | **1.14 (1.03, 1.26)** |
| | N | 2,053 | 3,329 | 2,306 | 750 | 340 | |
| | % | 3.5 | 3.6 | 3.8 | 4.8 | 6.5 | |
| Post-traumatic stress disorder | n | 189 | 292 | 143 | 50 | 19 | **1.10 (1.02, 1.19)** |
| | N | 1,440 | 1,472 | 586 | 350 | 135 | |
| | % | 13.1 | 19.8 | 24.4 | 14.3 | 14.0 | |
| Conduct disorder | n | 323 | 326 | 151 | 38 | 19 | 0.98 (0.92, 1.05) |
| | N | 3,105 | 2,213 | 1,480 | 265 | 295 | |
| | % | 10.4 | 14.7 | 10.2 | 14.3 | 6.4 | |
| Attention-deficit/hyperactivity disorder | n | 640 | 694 | 341 | 112 | 63 | *1.01 (0.95, 1.08) |
| | N | 1,699 | 2,416 | 996 | 313 | 31 | |
| | % | 37.7 | 28.7 | 34.2 | 35.8 | 206.5 | |
| Single disorder | n | 1,286 | 1,631 | 909 | 352 | 143 | **0.88 (0.85, 0.92)** |
| | N | 2,741 | 3,347 | 1,633 | 965 | 533 | |
| | % | 46.9 | 48.7 | 55.7 | 36.5 | 26.8 | |
| Multiple disorders | n | 253 | 298 | 128 | 35 | 23 | **0.84 (0.78, 0.90)** |
| | N | 3,600 | 4,577 | 2,916 | 817 | 470 | |
| | % | 7.0 | 6.5 | 4.4 | 4.3 | 4.9 | |

The table presents the number of 17-year-olds from South London in 2009-2024 who were diagnosed with each disorder (n), the number who were estimated to have met criteria for the disorder and used health services (N), and the rate of diagnosis provision in those estimated to have met criteria for the disorder and used health services (%; i.e., n/N as a percentage) during the past year, by Index of Multiple Deprivation 2015 (IMD) national quintile. The table also presents odds ratios (OR) and their 95% confidence intervals (95% CI) for associations between IMD national quintile and diagnosis provision in these young people estimated to have each disorder. Bold text indicates $p<0.05$. These confidence intervals (and p value indications) do not account for possible error of N in relation to the true number who met criteria for the disorder and used health services. *The number of 17-year-old South Londoners living in an IMD quintile 5 neighbourhood who had a diagnosis of ADHD was set at 31 (i.e., 100%) for this test to enable the analysis to run.

**X. Diagnosis provision in those estimated to have met criteria for a disorder and used health services, Sample 3, by ethnicity**

| | | White | Black, Mixed, Asian, or Other | White vs Black, Mixed, Asian, or Other OR (95% CI) |
|---|---|---|---|---|
| Any of the below disorders | n | 2,544 | 2,383 | **2.43 (2.27, 2.59)** |
| | N | 7,398 | 13,423 | |
| | % | 34.4 | 17.8 | |
| Depressive disorder | n | 1,010 | 983 | **2.04 (1.86, 2.25)** |
| | N | 5,906 | 10,724 | |
| | % | 17.1 | 9.2 | |
| Generalised anxiety disorder | n | 214 | 106 | **3.11 (2.45, 3.94)** |
| | N | 3,065 | 4,498 | |
| | % | 7.0 | 2.4 | |
| Post-traumatic stress disorder | n | 240 | 431 | **1.48 (1.25, 1.76)** |
| | N | 1,315 | 3,290 | |
| | % | 18.3 | 13.1 | |
| Conduct disorder | n | 439 | 425 | **2.69 (2.33, 3.10)** |
| | N | 2,454 | 5,669 | |
| | % | 17.9 | 7.5 | |
| Attention-deficit/hyperactivity disorder | n | 1,057 | 785 | **1.14 (1.00, 1.30)** |
| | N | 1,979 | 1,567 | |
| | % | 53.4 | 50.1 | |
| Single disorder | n | 2,143 | 2,047 | **5.98 (5.45, 6.56)** |
| | N | 3,062 | 7,294 | |
| | % | 70.0 | 28.1 | |
| Multiple disorders | n | 401 | 336 | **1.76 (1.51, 2.04)** |
| | N | 4,336 | 6,130 | |
| | % | 9.2 | 5.5 | |

The table presents the number of 17-year-olds from South London in 2009-2024 who were diagnosed with each disorder (n), the number who were estimated to have met criteria for the disorder and used health services (N), and the rate of diagnosis provision in those estimated to have met criteria for the disorder and used health services (%; i.e., n/N as a percentage) during the past year, by ethnicity. The table also presents odds ratios (OR) and their 95% confidence intervals (95% CI) for associations between ethnicity and diagnosis provision in these young people estimated to have each disorder. Bold text indicates $p<0.05$. These confidence intervals (and p value indications) do not account for possible error of N in relation to the true number who met criteria for the disorder and used health services.

### Y. Natural language processing pilot

Natural language processing (NLP) can be used to extract clinical information from free-text sections of electronic health records (EHRs). However, previous research has found that some NLP models demonstrate poor performance.[46–48] Additionally, external validation of clinical NLP models has been limited,[49] and there are concerns about poor generalisability when applied to new data,[50] including changes to cohort definitions within the same EHR system.[51] It is therefore recommended that NLP model performance is appropriately evaluated for each use and clearly reported. However, systematic reviews have found that reporting of NLP model evaluation is inconsistent, and sometimes not provided.[49,51]

We considered using the NLP application previously developed to identify mental health diagnoses in South London and Maudsley NHS Foundation Trust's EHR via Maudsley Biomedical Research Centre's Clinical Record Interactive Search.[52] This NLP application was developed based on adult records to identify a wide range of diagnoses, including post-traumatic stress disorder (PTSD). We evaluated its precision in identifying diagnoses in young people's records, focusing on PTSD. Our sample was all South London and Maudsley NHS Foundation Trust patients with an NLP PTSD diagnosis before age 18 years during an episode of care that started on or after 1st January 2008, when the EHR was used by all teams. Our data was extracted on 5th March 2026. We reviewed snippets of text (approximately 50-80 words) that contained the first NLP PTSD diagnosis for each patient, and determined whether the text truly indicated that the patient had a current PTSD diagnosis. If the snippet did not indicate that the patient had a current PTSD diagnosis, we reviewed the snippet of text containing the patient's second NLP PTSD diagnosis, and if that also did not indicate that the patient had a current PTSD diagnosis, we reviewed the snippet of text containing the patient's third NLP PTSD diagnosis. The researchers undertaking this review had received expert training about child and adolescent mental healthcare and PTSD, and comprehensive written and verbal guidance on this task. Initial and any unclear snippets were reviewed as a team with SJL (a Consultant Child and Adolescent Psychiatrist and specialist in PTSD) to achieve consensus.

We found poor precision of the NLP application in identifying PTSD diagnoses in this sample. Considering only each patient's first NLP PTSD diagnosis, we found that 52.1% (n=1,572/3,020) of patients had an accurate current PTSD diagnosis. Considering each patient's first three NLP PTSD diagnoses, we found that 58.2% (n=1,758/3,020) of patients had an accurate current PTSD diagnosis (any of the three). Common examples of the NLP application inaccurately identifying PTSD diagnoses involved text indicating family members as the experiencer (e.g. "the patient's mum has a PTSD diagnosis"), negation (e.g. "a PTSD diagnosis was ruled out"), possible diagnoses (e.g. "she has some PTSD symptoms and we are considering whether or not full criteria for diagnosis are met"), and historical temporality (e.g. "has had a PTSD diagnosis in the past"). These examples are not direct quotes, but reflect common themes.

We therefore concluded that NLP needs considerable development and evaluation before it can be confidently used to identify diagnoses like PTSD in young people's records in this data resource. Because PTSD is one of the key disorders we focus on in this study, and poor performance may well be present for other mental health disorder diagnoses in young people, we decided to use structured diagnoses only for the current analyses.

### Z. References

1 National Institute for Health and Care Excellence. Depression in children and young people: identification and management. 2019 www.nice.org.uk/guidance/ng134.

2 National Institute for Health and Care Excellence. Generalised anxiety disorder and panic disorder in adults: management. 2020 www.nice.org.uk/guidance/cg113.

3 Walter HJ, Bukstein OG, Abright AR, *et al.* Clinical Practice Guideline for the assessment and treatment of children and adolescents with anxiety disorders. *J Am Acad Child Adolesc Psychiatry* 2020; **59**: 1107–24.

4 National Institute for Health and Care Excellence. Post-traumatic stress disorder. 2018 www.nice.org.uk/guidance/ng116.

5 National Institute for Health and Care Excellence. Antisocial behaviour and conduct disorders in children and young people: recognition and management. National Institute for Health and Care Excellence, 2017 www.nice.org.uk/guidance/cg158.

6 National Institute for Health and Care Excellence. Attention deficit hyperactivity disorder: diagnosis and management. National Institute for Health and Care Excellence, 2019 www.nice.org.uk/guidance/ng87.

7 World Health Organization. The ICD-10 classification of mental and behavioural disorders. Geneva: World Health Organization, 1992.

8 American Psychiatric Association. Diagnostic and Statistical Manual of Mental Disorders, 4th edition. Washington DC: American Psychiatric Association, 1994.

9 Robins LN, Cottler L, Bucholz KK, Compton W. Diagnostic Interview Schedule for DSM-IV. St Louis, MO: Washington University School of Medicine, 1995.

10 Moffitt TE, Caspi A, Taylor A, *et al.* How common are common mental disorders? Evidence that lifetime prevalence rates are doubled by prospective versus retrospective ascertainment. *Psychol Med* 2010; **40**: 899–909.

11 American Psychiatric Association. Diagnostic and Statistical Manual of Mental Disorders, 5th edition. Washington DC: American Psychiatric Association, 2013.

12 Lewis SJ, Arseneault L, Caspi A, *et al.* The epidemiology of trauma and post-traumatic stress disorder in a representative cohort of young people in England and Wales. *Lancet Psychiatry* 2019; **6**: 247–56.

13 Agnew-Blais JC, Polanczyk GV, Danese A, Wertz J, Moffitt TE, Arseneault L. Evaluation of the persistence, remission, and emergence of attention-deficit/hyperactivity disorder in young adulthood. *JAMA Psychiatry* 2016; **73**: 713.

14 Stewart R, Soremekun M, Perera G, *et al.* The South London and Maudsley NHS Foundation Trust Biomedical Research Centre (SLAM BRC) case register: development and descriptive data. *BMC Psychiatry* 2009; **9**: 51.

15 Fernandes AC, Cloete D, Broadbent MT, *et al.* Development and evaluation of a de-identification procedure for a case register sourced from mental health electronic records. *BMC Med Inform Decis Mak* 2013; **13**: 71.

16 Perera G, Broadbent M, Callard F, *et al.* Cohort profile of the South London and Maudsley NHS Foundation Trust Biomedical Research Centre (SLaM BRC) case register: current status and recent enhancement of an electronic mental health record-derived data resource. *BMJ Open* 2016; **6**: e008721.

17 UK Ministry of Housing, Communities and Local Government. English indices of deprivation 2015. 2015 www.gov.uk/government/statistics/english-indices-of-deprivation-2015.

18 UK Ministry of Housing, Communities and Local Government. English indices of deprivation 2019. 2019 www.gov.uk/government/statistics/english-indices-of-deprivation-2019.

19 UK Ministry of Housing, Communities and Local Government. English indices of deprivation 2025. 2025 www.gov.uk/government/statistics/english-indices-of-deprivation-2025.

20 UK Office for National Statistics. Lower layer super output area population estimates. 2025 www.ons.gov.uk/peoplepopulationandcommunity/populationandmigration/populationestimates/datasets/lowersuperoutputareamidyearpopulationestimates.

21 UK Office for National Statistics. Census 2011: Ethnic group by sex by age, DC2101EW. 2013 www.nomisweb.co.uk/census/2011/dc2101ew.

22 UK Office for National Statistics. Census 2021: Age, ethnic group and sex, by local authority. 2024 www.ons.gov.uk/datasets/create/filter-outputs/4e9c43fe-376a-4755-bdb8-be63f1087866#get-data.

23 Kessler RC, Wittchen H-U, Abelson JM, *et al.* Methodological studies of the Composite International Diagnostic Interview (CIDI) in the US national comorbidity survey (NCS). *Int J Methods Psychiatr Res* 1998; **7**: 33–55.

24 Kessler RC, Avenevoli S, Green J, *et al.* National Comorbidity Survey Replication Adolescent Supplement (NCS-A): III. Concordance of DSM-IV/CIDI diagnoses with clinical reassessments. *J Am Acad Child Adolesc Psychiatry* 2009; **48**: 386–99.

25 Haro JM, Arbabzadeh-Bouchez S, Brugha TS, *et al.* Concordance of the Composite International Diagnostic Interview Version 3.0 (CIDI 3.0) with standardized clinical assessments in the WHO World Mental Health Surveys. *Int J Methods Psychiatr Res* 2006; **15**: 167–80.

26 Eaton WW, Neufeld K, Chen L-S, Cai G. A comparison of self-report and clinical diagnostic interviews for depression: diagnostic interview schedule and schedules for clinical assessment in neuropsychiatry in the Baltimore Epidemiologic Catchment Area follow-up. *Arch Gen Psychiatry* 2000; **57**: 217.

27 Murphy JM, Monson RR, Laird NM, Sobol AM, Leighton AH. A comparison of diagnostic interviews for depression in the Stirling County Study: challenges for psychiatric epidemiology. *Arch Gen Psychiatry* 2000; **57**: 230.

28 Kessler RC, Little RJA, Groves RM. Advances in strategies for minimizing and adjusting for survey nonresponse. *Epidemiol Rev* 1995; **17**: 192–204.

29 De Los Reyes A, Augenstein TM, Wang M, *et al.* The validity of the multi-informant approach to assessing child and adolescent mental health. *Psychol Bull* 2015; **141**: 858–900.

30 Jack RH, Joseph RM, Hollis C, *et al.* Seasonal trends in antidepressant prescribing, depression, anxiety and self-harm in adolescents and young adults: an open cohort study using English primary care data. *BMJ Ment Health* 2023; **26**: e300855.

31 Catalogue of Mental Health Measures. 2026 www.cataloguementalhealth.ac.uk.

32 Moffitt TE, the E-Risk Study Team. Teen-aged mothers in contemporary Britain. *J Child Psychol Psychiatry* 2002; **43**: 727–42.

33 Gjone H, Nøvik TS. Parental ratings of behaviour problems: a twin and general population comparison. *J Child Psychol Psychiatry* 1995; **36**: 1213–24.

34 Kendler KS, Martin NG, Heath AC, Eaves LJ. Self-report psychiatric symptoms in twins and their nontwin relatives: are twins different? *Am J Med Genet* 1995; **60**: 588–91.

35 Moilanen I, Linna SL, Ebeling H, *et al.* Are twins' behavioural/emotional problems different from singletons'? *Eur Child Adolesc Psychiatry* 1999; **8**: S62–7.

36 Schumann G, Loth E, Banaschewski T, *et al.* The IMAGEN study: reinforcement-related behaviour in normal brain function and psychopathology. *Mol Psychiatry* 2010; **15**: 1128–39.

37 Maričić LM, Walter H, Rosenthal A, *et al.* The IMAGEN study: a decade of imaging genetics in adolescents. *Mol Psychiatry* 2020; **25**: 2648–71.

38 Vizard T, Sadler K, Ford T, *et al.* Mental Health of Children and Young People in England, 2017: survey design and methods report. NHS Digital, 2018 https://digital.nhs.uk/data-and-information/publications/statistical/mental-health-of-children-and-young-people-in-england/2017/2017.

39 Knowles G, Gayer-Anderson C, Blakey R, *et al.* Cohort Profile: Resilience, Ethnicity and AdolesCent mental Health (REACH). *Int J Epidemiol* 2022; **51**: e303–13.

40 Trouton A, Spinath FM, Plomin R. Twins early development study (TEDS): a multivariate, longitudinal genetic investigation of language, cognition and behavior problems in childhood. *Twin Res* 2002; **5**: 444–8.

41 Odgers CL, Caspi A, Russell MA, Sampson RJ, Arseneault L, Moffitt TE. Supportive parenting mediates neighborhood socioeconomic disparities in children's antisocial behavior from ages 5 to 12. *Dev Psychopathol* 2012; **24**: 705–21.

42 UK Office for National Statistics. Census 2011: Ethnic group by sex by age, LC2101EW. 2013 www.nomisweb.co.uk/census/2011/lc2101ew.

43 UK Office for National Statistics. Census 2021: Age, ethnic group and sex, by LSOA. 2023 www.ons.gov.uk/datasets/create/filter-outputs/ccd97b14-8ae6-47b9-a7c7-752351471518#get-data.

44 StataCorp. Stata statistical software: release 18. 2023.

45 R Core Team. R: a language and environment for statistical computing. 2026. www.R-project.org.

46 Dahl S, Bøgsted M, Sagi T, Vesteghem C. Performance of natural language processing for information extraction from electronic health records within cancer: systematic review. *JMIR Med Inform* 2025; **13**: e68707–e68707.

47 Hutto A, Zikry TM, Bohac B, *et al.* Using a natural language processing toolkit to classify electronic health records by psychiatric diagnosis. *Health Informatics J* 2024; **30**: 14604582241296411.

48 Rijcken E, Zervanou K, Mosteiro P, Scheepers F, Spruit M, Kaymak U. Machine learning vs. rule-based methods for document classification of electronic health records within mental health care—a systematic literature review. *Nat Lang Process J* 2025; **10**: 100129.

49 Kersloot MG, Van Putten FJP, Abu-Hanna A, Cornet R, Arts DL. Natural language processing algorithms for mapping clinical text fragments onto ontology concepts: a systematic review and recommendations for future studies. *J Biomed Semant* 2020; **11**: 14.

50 Kühnel L, Fluck J. We are not ready yet: limitations of state-of-the-art disease named entity recognizers. *J Biomed Semant* 2022; **13**: 26.

51 Fu S, Wang L, Moon S, *et al.* Recommended practices and ethical considerations for natural language processing-assisted observational research: a scoping review. *Clin Transl Sci* 2023; **16**: 398–411.

52 NIHR Maudsley Biomedical Research Centre. CRIS NLP applications library: diagnosis. 2026 www.maudsleybrc.nihr.ac.uk/facilities/clinical-record-interactive-search-cris/natural-language-processing-nlp-service/cris-nlp-applications-library/#Diagnosis.
